# Automated Segmentation of Intracranial Arteries on 4D Flow MRI for Hemodynamic Quantification

**DOI:** 10.64898/2026.03.09.26347567

**Authors:** Jiaxin Zhang, Anouk S. Verschuur, Pim van Ooij, Eric M. Schrauben, Mark Bakker, Kyung Min Nam, Irene C. van der Schaaf, Chantal M.W. Tax

## Abstract

Accurate vessel segmentation is essential for reliable hemodynamic quantification in 4D Flow MRI. Automated segmentation with deep learning offers a promising alternative to the time-consuming, operator-dependent manual segmentation, but its application is often hindered by the scarcity of labeled datasets. Moreover, while a previous study has evaluated the impact of automated vs manual segmentation on downstream intracranial hemodynamic quantification, it remains to be explored whether variability between different automated segmentation methods affects hemodynamic quantification significantly, particularly in the context of intracranial 7T 4D Flow MRI. We investigated a transfer learning–based intracranial artery segmentation model using a 3D full-resolution nnU-Net, pretrained on 355 TOF-MRA scans and fine-tuned on 11 7T 4D Flow MRI scans. The model was compared with two published models: a direct-transfer U-Net pretrained only on TOF-MRA without 4D Flow MRI fine-tuning, and a DenseNet U-Net trained from scratch on the same 11 7T 4D Flow MRI scans. Segmentation performance was evaluated against the manual reference on test sets with different resolutions and the impact on downstream hemodynamic quantification was assessed. The transfer learning–based nnU-Net achieved the highest Dice score (>0.85), the lowest HD95 (∼3 mm), and the highest ICCs in cross-sectional area (0.62–0.87, except PCAs) and mean blood flow (0.78– 0.98). For wall shear stress (WSS) quantification, the transfer learning–based nnU-Net segmentations achieved the closest agreement with the manual reference (mean = 1.57 ± 0.63 Pa, ICC = 0.96; max = 2.16 ± 1.05 Pa, ICC = 0.97) and minimal bias (≤ 1.7%), whereas U-Net and DenseNet U-Net showed systematic under-(−5%) and overestimation (+7%), respectively. However, several vessel segments, including the ACA for DenseNet U-Net and the BA for U-Net, showed statistically significant differences (ANOVA post-hoc correction P < 0.05) in the flow-related metrics when compared with the manual reference. These results suggest that a transfer learning–based nnU-Net can provide accurate automated intracranial artery segmentation for 7T 4D Flow MRI. Differences among automated segmentation pipelines and associated analysis regions were linked to differences in downstream hemodynamic estimates.

## 1 Introduction

Hemodynamics plays a crucial role in the onset and progression of cerebrovascular diseases. Accurate and in vivo real-time assessment of hemodynamics is essential for improving early prediction and treatment planning in patients with cerebrovascular diseases (Tuijl et al., 2024). 4D Flow MRI is an advanced non-invasive imaging modality that enables comprehensive, time-resolved quantification of three-dimensional blood flow, where ultra-high field strengths such as 7 T can provide higher signal-to-noise ratio (SNR) and image resolution for detailed characterization of intracranial vasculature (Markl et al., 2012; van Ooij, Zwanenburg, et al., 2013).

A prerequisite for accurate hemodynamics assessment is the precise and robust segmentation of intracranial vessels (Dunås et al., 2019). Manual segmentation of the intracranial vessels is time-consuming and also prone to inter-observer variability, which can affect the reproducibility of clinical measurements (Rothenberger et al., 2023). While semi- or fully automated segmentation methods have been proposed recently, they often still require manual intervention or lack specific focus on intracranial arteries (Rothenberger et al., 2023; Schrauben et al., 2015).

Recently, deep learning (DL) methods, particularly convolutional neural networks (CNNs), have shown great promise for the segmentation of intracranial vessels (de Vos et al., 2021; Sobisch et al., 2022). In particular, nnU-Net is a self-configuring CNN-based framework that automates the entire segmentation process, and has shown strong performance across diverse medical image segmentation tasks (Isensee et al., 2021). However, most DL studies primarily focus on imaging techniques such as time-of-flight magnetic resonance angiography (TOF-MRA), computed tomography angiography (CTA) and digital subtraction angiography (DSA) (Kim et al., 2025; Min et al., 2025; Su et al., 2024), with only one study exploring on 4D flow MRI to our knowledge (Winter et al., 2024). Furthermore, as 4D Flow MRI at 7T has not yet been widely implemented in clinical practice, obtaining sufficient data to train large-scale networks from scratch remains challenging.

To address this gap, we adopt a transfer learning–based intracranial artery segmentation strategy for 4D Flow MRI, built upon the nnU-Net framework. This model was pre-trained on a large dataset of brain TOF-MRA images and fine-tuned on a smaller 4D Flow MRI dataset. The fine-tuned model was then evaluated against the manual reference and compared with two previously published intracranial artery segmentation models, one pretrained on TOF-MRA and the other trained from scratch using the same 4D Flow MRI dataset used for fine-tuning, to assess segmentation performance across different acquisition protocols and spatial resolutions. Furthermore, we evaluated how the different segmentation methods affect the quantification of 4D Flow MRI–derived hemodynamic parameters, providing insights into the extent of variability introduced by segmentation differences.

## 2 Method

### 2.1 Study cohort

Our study cohort was composed of two datasets containing TOF-MRA and 4D flow MRI scans. The TOF-MRA scans were derived from the publicly available CerebrOvascular SegmenTAtion dataset (COSTA) which comprises 355 high-quality annotated TOF-MRA images from six different data centers (Mou et al., 2023). This multi-center, multi-vendor dataset includes scans acquired on both 1.5T and 3T MRI systems (Philips, GE, Siemens, and United Imaging Healthcare) from healthy subjects and patients with IAs or Moyamoya disease. The voxel spacing varied across scans, with a median in-plane spacing of 0.47 × 0.47 mm and a median slice spacing of 0.80 mm. The 4D Flow MRI scans were acquired from two local 7T 4D Flow MRI studies performed at the University Medical Center Utrecht (UMCU), the Netherlands. The first was the Flow@Aneurysm study, which enrolled 37 patients (June 2021 - July 2022) with unruptured intracranial aneurysms (IAs) larger than 4 mm (Tuijl et al., 2024; Van Tuijl et al., 2024). The second was the DETAILING study, which prospectively recruited 3 patients with unruptured IAs (May 2024 - February 2025). In addition, 2 healthy volunteers were scanned for workflow testing prior to patient data acquisition (May 2024 – September 2024). Both studies were approved by the local ethics review committee.

### 2.2 4D flow MRI data

7T MRI data were acquired using a Philips Healthcare scanner (Best, The Netherlands) equipped with an 8-channel transmit and a 32-channel receive coil (Nova Medical, Houston, USA). Both studies utilized the "PROspective Undersampling in multiple Dimension" (PROUD) patch for acceleration, with the DETAILING study using an updated software version of the same scanner (Gottwald, Peper, et al., 2020). All scans covered the Circle of Willis (CoW) to ensure inclusion of IAs, with retrospective gating performed using a peripheral pulse unit. The acquisition duration for all scans was approximately 10 minutes. The acquisition parameters for both studies are detailed in Supplementary Table 1. Both low-resolution (near-isotropic, ∼0.7 mm) and high-resolution (isotropic, 0.5 mm) 4D flow MRI scans were acquired, with VENC values of 100 cm/s and 50 cm/s, respectively, to accommodate the range of expected flow velocities within aneurysms and the CoW in general.

All 4D flow MRI data were reconstructed using an in-house MATLAB pipeline developed by Amsterdam UMC for the PROUD patch (Gottwald, Peper, et al., 2020), which integrated the Berkeley Advanced Reconstruction Toolbox (BART) and ReconFrame (Gyrotools, Zurich, CH) (Uecker et al., 2016). Phase corrections were applied during 4D Flow MRI reconstruction. Concomitant gradient field correction was performed to reduce spatially varying phase errors caused by Maxwell terms using the MRecon ConcomitantFieldCorrection method with scanner-provided correction parameters. Residual background phase offsets caused by eddy currents were corrected using the MRecon FlowPhaseCorrection method, in which a third-order polynomial phase field was estimated from static tissue regions and subtracted from the phase images (Busch et al., 2017). The FlowPhaseCorrection step was applied using the default MRecon implementation, without user-defined parameters for static-tissue selection or low-SNR voxel exclusion. No explicit phase unwrapping or velocity-aliasing correction was performed.

After reconstruction, all 4D Flow MRI datasets underwent visual quality control by one researcher (JZ). Motion artifacts were graded using a clinical motion artifact scale, ranging from none, minimal, and mild to moderate and severe (Andre et al., 2015). In this study, scans with moderate or severe motion artifacts, corresponding to marginal diagnostic quality or non-diagnostic quality, were excluded because reliable manual vessel segmentation was not possible. Scans acquired in sagittal orientation were also excluded. Finally, 11 low-resolution scans from 11 patients in the Flow@Aneurysm study were randomly selected and are hereafter to as the fine-tuning set. The remaining scans from both studies, comprising 19 low-resolution and 13 high-resolution scans from 22 patients and 2 healthy volunteers, were used as separate low- and high-resolution test sets. No patient included in the low-resolution fine-tuning set was represented in either of the low-resolution or high-resolution test cohorts.

### 2.3 Postprocessing and manual reference generation

Phase Contrast Magnetic Resonance Angiography (PCMRA) images generated from the 4D flow MRI data served as the foundation of our segmentation study. Because PCMRA highlights blood vessels with flow while suppressing static regions, it is well suited for vascular segmentation (Hennemuth et al., 2011). PCMRA images were computed by a time-averaged combination of the magnitude image (M) and the velocity components (Vx, Vy, Vz) over the entire cardiac cycle, as follows (Köhler et al., 2017):

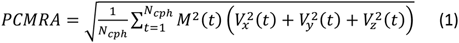

where N_cph_ is the number of cardiac phases. No explicit noise mask was applied before PCMRA calculation.

For the generation of the manual reference segmentation of the CoW, the PCMRA images were segmented in 3D Slicer (version 5.6.2). The segmentation workflow involved an initial step of thresholding and region-growing, followed by manual corrections. The reference segmentations used for model training and evaluation were performed by JZ, who was trained by the interventional neuroradiologist (ICvdS; over 20 years of experience. To assess inter-observer variability, 8 test scans, corresponding to approximately 25% of the test cases, were randomly selected, including 4 low-resolution and 4 high-resolution scans. A second observer (ICvdS) segmented these cases in 3D Slicer after being informed of the anatomical range to be segmented, without access to the first observer’s reference segmentation and the automated model outputs.

### 2.4 Transfer learning model

For intracranial artery segmentation on 4D Flow MRI using transfer learning, we employed a 3D full-resolution nnU-Net v2 configuration (Isensee et al., 2021). We trained this model from scratch using 355 TOF-MRA image and segmentation pairs from the COSTA dataset. To maximize the data available for transfer learning, pre-training was performed using all images by setting the nnU-Net fold configuration to “all”, i.e., without a separate validation set, for 1000 epochs with an initial learning rate of 0.01. To adapt this pre-trained model on TOF-MRA to the 4D Flow MRI domain, fine-tuning was performed using the dedicated fine-tuning set, with 5-fold cross-validation, an initial learning rate of 0.001, and 100 epochs for each fold. The nnU-Net framework automatically handled the data pre-processing for both pre-training and fine-tuning, including resampling to a unified voxel spacing and Z-score normalization. The target voxel spacing was automatically determined from the 4D Flow MRI fine-tuning set as 0.70 × 0.66 × 0.66 mm and was applied consistently during pre-training, fine-tuning and inference. The patch size was 32 × 256 × 256, with a batch size of 2 for the 3D full-resolution configuration. During inference, the default nnU-Net inference strategy was used, including sliding-window prediction, Gaussian-weighted tile aggregation, and test-time mirroring. The 5-fold-specific nnU-Net models obtained during cross-validation were ensembled by averaging the predicted class probabilities before conversion to the final segmentation. The flowchart for the transfer learning model is shown in Figure 1. For clarity, the transfer-learning model is hereafter referred to as the fine-tuned nnU-Net throughout the manuscript.

**Figure 1:**
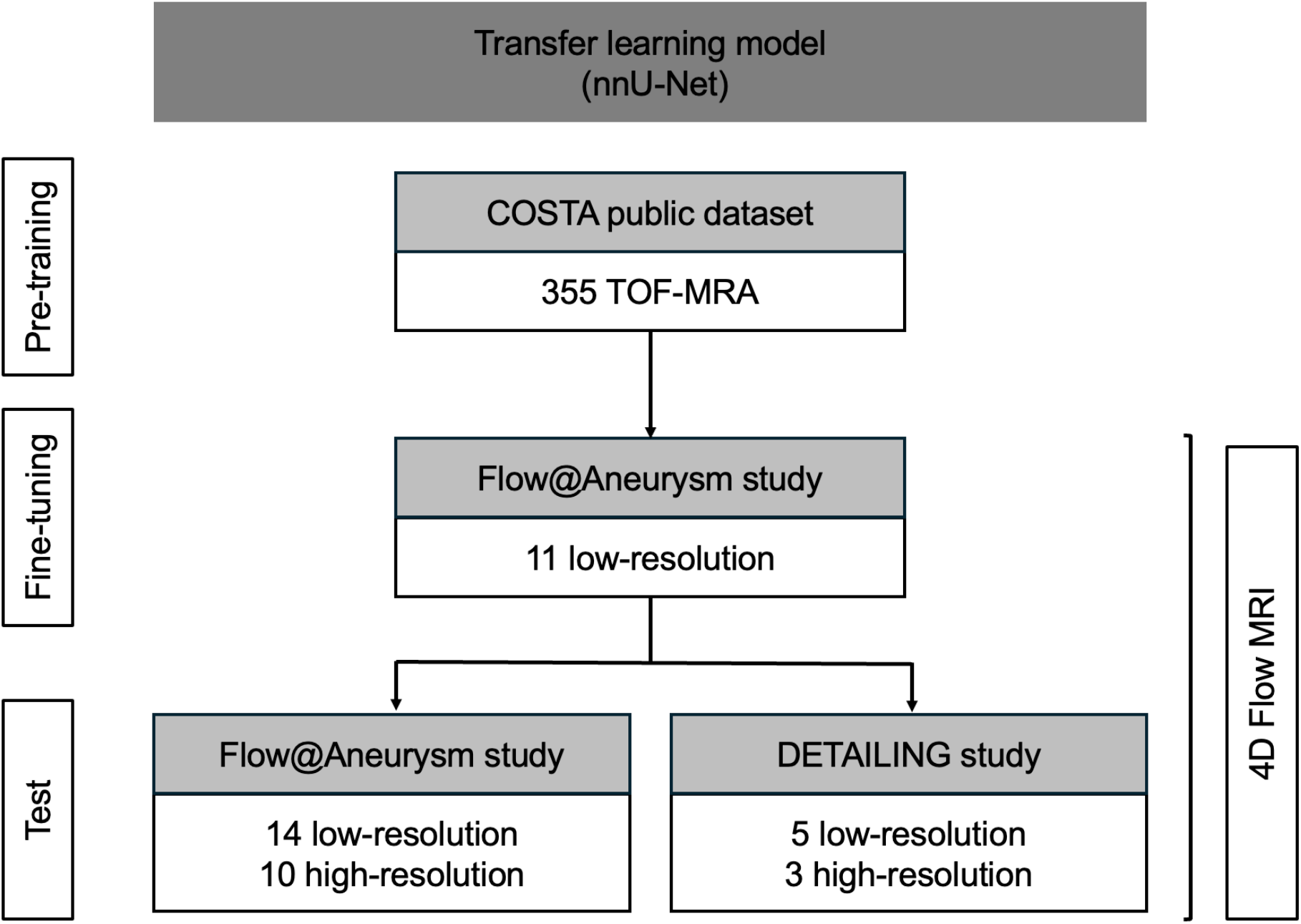
Data flowchart of the transfer learning segmentation model from TOF-MRA to 4D flow MRI using nnU-Net.

To evaluate the contribution of TOF-MRA pre-training and 4D Flow MRI fine-tuning, we performed additional nnU-Net ablation experiments. The TOF-MRA-pretrained nnU-Net applied directly to 4D Flow MRI without fine-tuning is hereafter referred to as the direct-transfer nnU-Net. In the full-data setting, nnU-Net was also trained from scratch using the same fine-tuning set with 5-fold cross-validation for 1000 epochs and an initial learning rate of 0.01. All other nnU-Net settings were kept the same as in the fine-tuned nnU-Net. To assess robustness under limited annotation availability, fine-tuned and from-scratch nnU-Net models were compared using reduced fine-tuning subsets of 1, 3, 5, 7, 9, and 11 cases. These subsets formed a single nested subset sequence, generated by randomly removing two cases at each step. The number of folds was adjusted according to the number of available cases: 1 fold was used for the one-case setting, 3 folds for the three-case setting, and 5 folds for settings with five or more cases. For the from-scratch models, both 200-epoch and 1000-epoch training schedules were evaluated. Unless otherwise specified, the full-data 1000-epoch model is referred to as the from-scratch nnU-Net.

### 2.5 Comparative segmentation models

Our study employed two segmentation models from previous studies for comparison, both based on the U-Net architecture.

The first model, hereafter referred to as U-Net, was a pre-trained (n=69) 3D U-Net for cerebral vessel segmentation on TOF-MRA data (de Vos et al., 2021). For data pre-processing, we applied Z-score normalization and padded the images to ensure they could be divided into 64×64×64 patches. We used the provided pre-trained weights and ran inference on our 4D Flow MRI test sets without additional fine-tuning.

The second model, referred to as DenseNet U-Net, was based on the CNN architecture originally proposed by Berhane et al. (Berhane et al., 2020) and later applied to intracranial vessel segmentation on 4D Flow MRI by Winter et al. (Winter et al., 2024). Because the trained weights from the previous intracranial 4D Flow MRI study were not publicly available, the DenseNet U-Net was trained from scratch on our 4D Flow MRI fine-tuning set. The implementation was adapted from the open-source TensorFlow 1.x code provided by Winter et al. (https://github.com/drsol1986/AI-Segmentation-) and re-implemented in PyTorch to ensure compatibility with the current hardware and software environment, including newer CUDA versions. The core network design, including the dense-block structure, was preserved as closely as possible during this adaptation. For data pre-processing, images were first rescaled using min-max normalization and then center-cropped or padded to a fixed dimension of 224×192×64. This model was trained on the fine-tuning set using 5-fold cross-validation, with a batch size of 1, a learning rate of 0.0001, and 100 epochs for each fold. For inference, the 5-fold-specific models were ensembled using voxel-wise majority voting on the binary predictions.

### 2.6 Segmentation performance analysis

All models trained in this study, including the DenseNet U-Net and nnU-Net variants, were trained on a server equipped with an NVIDIA RTX A5000 GPU, using PyTorch 2.8.0 with CUDA 12.8. All segmentation models were evaluated against the manual reference segmentation on both the low-resolution and high-resolution test sets. We used two widely adopted metrics: Dice Score (DS) and the 95th-percentile Hausdorff Distance (HD95). The DS quantifies the volumetric overlap between the predicted segmentation and the reference and ranges from 0 to 1, with higher values indicating better overlap. HD95 assesses boundary accuracy and is computed as the 95^th^ percentile of the bidirectional Euclidean distances between the boundary points of the predicted segmentation and the reference. A smaller HD95 value signifies closer spatial alignment and higher boundary fidelity.

For the main segmentation comparison, involving the direct-transfer U-Net, DenseNet U-Net, and fine-tuned nnU-Net, we adopted a multi-stage evaluation. First, we compared each model’s segmentation with the manual reference to reflect native performance. Second, because the U-Net model was not specifically trained for CoW segmentation and segmented intracranial vessels more broadly, we introduced a cropping step to standardize the region of interest (ROI). A bounding box was defined around the manual reference CoW in 3D Slicer by JZ and was applied to the segmentations from the three main models using a custom Python script to compute the performance metrics against the manual reference. Finally, since the cropped segmentations from the U-Net and DenseNet U-Net models still contained small non-CoW branches in some subjects, we performed manual cleaning on these outputs to recompute the metrics against the manual reference. In contrast, most irrelevant vascular structures in the fine-tuned nnU-Net segmentations had already been removed during the cropping step, and therefore no additional manual cleaning was performed for the cropped fine-tuned nnU-Net outputs.

### 2.7 Hemodynamic analysis

All hemodynamic analyses were performed for the three main segmentation models, namely the U-Net, DenseNet U-Net, and fine-tuned nnU-Net, using the manual reference segmentation as the reference. The high-resolution 4D Flow MRI data (VENC = 50 cm/s) were excluded from hemodynamic analysis because velocity aliasing was observed in some scans and no explicit phase-unwrapping or velocity-unaliasing correction was performed.

#### 2.7.1 Flow analysis

Flow analysis was performed using a modified version of the open-source Quantitative Velocity Tool (QVT) software, which is publicly available at https://github.com/uwmri/QVT (Roberts et al., 2023; Schrauben et al., 2015). We modified the QVT to directly import 3D segmentation masks from the U-Net, DenseNet U-Net, fine-tuned nnU-Net and the manual reference, bypassing its built-in algorithm for global vessel segmentation. Based on these masks, QVT then automatically generated vessel centerlines and placed orthogonal analysis planes along them to enable quantification.

The analysis planes were placed at key anatomical landmarks within the CoW. Specifically, planes were positioned on the mid-basilar artery (BA), as well as halfway along the A1 segment of the anterior cerebral artery (ACA), the M1 segment of the middle cerebral artery (MCA), the P1 segment of the posterior cerebral artery (PCA), and the cavernous segment of the internal carotid artery (ICA) (Supplementary Fig.1). Based on these analysis planes, the time-constant cross-sectional arterial area, as well as the mean blood flow and mean velocity averaged across the entire cardiac cycle, were automatically quantified by QVT.

#### 2.7.2 Wall shear stress (WSS) estimation

WSS was computed from the 4D Flow MRI velocity field using an in-house MATLAB-based framework from previous work (van Ooij, Potters, et al., 2013). Briefly, vessel surfaces were extracted from the segmentation masks and smoothed using Laplacian filtering with 15 iterations and a step size of λ = 0.1. For each surface vertex, the local radius was estimated from the nearest intersection with the opposite vessel surface along the inward wall-normal direction. Velocity values were then sampled along the inward wall-normal direction at three equidistant points, corresponding to one-third, two-thirds, and the full local radius from the vessel wall. Velocity values at off-grid sampling locations were obtained using natural-neighbor interpolation based on a Delaunay triangulation of the velocity grid, with the vessel wall assigned zero velocity to impose a no-slip boundary condition. After rotating the local coordinate system to align the wall normal with the z-axis, the tangential velocity components were fitted with smoothing splines along the normal direction with weights of [0.1, 1, 1, 1] and an adaptive tolerance of max(|vₜ|)/100, where vₜ denotes the sampled tangential velocity profile. WSS was then calculated as the product of blood viscosity (3.5 × 10^-3^ Pa·s) and the tangential velocity gradient at the vessel wall, and the Euclidean norm of the WSS vector was used for subsequent analysis. WSS values were computed for each cardiac phase using the same time-averaged segmentation mask. For each phase, WSS values were spatially aggregated over the entire CoW surface by taking the arithmetic mean over non-zero WSS surface vertices. We then evaluated the temporal mean (mean WSS) and temporal maximum (max WSS) of these values across the entire cardiac cycle. Additionally, WSS visualizations were generated for a representative case at the systolic cardiac phase to provide qualitative insights. To ensure consistency in the WSS calculation region, the analysis was performed on the cropped segmentation masks, with the manual cleaning for the U-Net and DenseNet U-Net models (as described in 2.6).

### 2.8 Statistical analysis

We first assessed normality of all hemodynamic parameters (arterial area, mean flow and mean velocity for each arterial segment; mean WSS, and max WSS for the whole CoW) using the Shapiro–Wilk test. Given the robustness of repeated-measures analysis of variance (ANOVA) to mild deviations from normality, and for consistency across the hemodynamic analyses, one-way repeated-measures ANOVA was applied consistently across all hemodynamic comparisons (factor: method; levels: U-Net, DenseNet U-Net, fine-tuned nnU-Net, manual reference). The Greenhouse–Geisser correction was applied to account for potential violations of sphericity. Post-hoc comparisons with a Bonferroni adjustment were performed for all pairwise comparisons. Agreement and reproducibility were quantified using the intraclass correlation coefficients (ICCs) and Bland–Altman analysis. ICCs with 95% confidence intervals were calculated between each automated method and the manual reference using a two-way mixed-effects model with absolute agreement and single measurement. For Bland–Altman, we used percent differences defined as 100 × (Method − Reference)/[(Method + Reference)/2], and report bias ± 95% limits of agreement (LOA) in percent. ICCs were calculated using the Reliability Analysis procedure in IBM SPSS Statistics, version 25.0 (IBM Corp., Armonk, NY, USA); all other analyses were performed using GraphPad Prism, version 10.4.1 (GraphPad Software, Boston, MA, USA). A two-sided P value < 0.05 (after Bonferroni adjustment where applicable) was considered statistically significant.

## 3 Results

### 3.1 Baseline Characteristics

In the 37 patients from the Flow@Aneurysm 4D Flow MRI study, 2 patients were excluded due to sagittal acquisition and 5 patients were excluded due to moderate or severe motion artifacts. No obvious velocity wrapping artifacts were observed in the low-resolution datasets used for hemodynamic quantification during visual inspection. In the high-resolution datasets, velocity wrapping was observed in some scans, which were retained because they were not used for hemodynamic quantification. After quality control, hypoplastic vessel segments and segments that could not be reliably analyzed using QVT were excluded. The latter included segments with incomplete or discontinuous centerlines, unreliable placement of analysis planes, or inconsistent local flow estimates. The same vessel segments were retained across all four methods. This resulted in ACA (R/L = 8/11), MCA (14/12), PCA (8/8), ICA (17/15), and BA (15) segments being included for subsequent blood flow analysis in QVT.

The characteristics of the included subjects and scans are summarized in Table 1.

**Table 1:** Overview of cohort characteristics and 4D Flow MRI scan inclusion.

|  | **Flow@Aneurysm study** | **DETAILING study** | **Total** |
|---|---|---|---|
| Total subjects | 37 patients | 3 patients + 2 volunteers | 42 |
| Included subjects for segmentation | 30 patients | 3 patients + 2 volunteers | 35 |
| Women | 23 (77%) | 1 (20%) | 24 |
| Age, year ± SD | 62 ± 13 | 43 ± 18 | 59 ± 15 |
| Low-resolution scans | 25 patients | 3 patients + 2 volunteers | 30 |
| High-resolution scans | 10 patients | 2 patients + 1 volunteer | 13 |

**Note**: The total number of subjects is 42. Some patients had both low- and high-resolution scans.

### 3.2 CoW segmentation performance

#### 3.2.1 Main model comparison

Table 2 reports on segmentation performance of the three main models at different evaluation stages (non-edited output, after cropping, and after cropping plus manual cleaning) compared with the manual reference under the training, domain-adaptation, and preprocessing conditions evaluated in this study. At the first stage (non-edited output), the fine-tuned nnU-Net pipeline achieved higher segmentation performance than the evaluated U-Net and DenseNet U-Net pipelines both in the high and lower resolution 4D flow MRI scans (fine-tuned nnU-net low resolution: DS = 0.860, HD95 = 2.98 mm; high resolution: DS = 0.856, HD95 = 3.12 mm). In the second stage (after cube cropping), all three models showed a general improvement in performance metrics with the largest gains observed for U-Net (low resolution: + 0.232 DS, -46.04 mm HD95; high resolution: + 0.250 DS, -66.23 mm HD95), while DenseNet U-Net showed modest changes. Fine-tuned nnU-Net showed the best performance in low-resolution data and similar performance to U-Net in high-resolution data (low resolution: DS = 0.876, HD95 = 0.99 mm; high resolution: DS = 0.863, HD95 = 1.20 mm). A final manual-cleaning step, which was applied only to U-Net and DenseNet U-Net, yielded further improvement for U-Net on low-resolution data (DS = 0.848, HD95 = 1.74 mm), while having a minimal effect on both models’ high-resolution data. Overall, the fine-tuned nnU-Net at the cropping stage (i.e., without manual cleaning) had overall best segmentation performance.

**Table 2.** Segmentation performance metrics for three models at different evaluation stages. HD95 indicates the 95% Hausdorff distance. All values are presented as mean [range].

|  |  | Non-edited output |  |  | Cube cropping |  |  | Cube cropping<br>+ manual cleaning* |  |
|---|---|---|---|---|---|---|---|---|---|
|  |  | **U-Net** | **DenseNet U-Net** | **Fine-tuned nnU-Net** | **U-Net** | **DenseNet U-Net** | **Fine-tuned nnU-Net** | **U-Net** | **DenseNet U-Net** |
| Dice score | Low resolution | 0.578 [0.457-0.695] | 0.813 [0.649-0.919] | **0.860** [0.732-0.946] | 0.810 [0.715-0.902] | 0.825 [0.649-0.932] | **0.876** [0.777–0.949] | **0.848** [0.771-0.910] | 0.828 [0.649-0.935] |
|  | High resolution | 0.618 [0.389-0.756] | 0.732 [0.270-0.869] | **0.856** [0.739-0.927] | **0.868** [0.798-0.929] | 0.734 [0.270-0.870] | 0.863 [0.773-0.927] | **0.869** [0.798-0.929] | 0.734 [0.270-0.870] |
| HD95 (mm) | Low resolution | 51.79 [31.58-81.66] | 5.06 [0.66-14.07] | **2.98** [0.66-28.04] | 5.75 [0.66-13.75] | 3.58 [0.66-14.07] | **0.99** [0.66-3.62] | **1.74** [0.66-8.79] | 3.54 [0.66-14.07] |
|  | High resolution | 67.38 [47.95-96.84] | 4.95 [0.50-22.94] | **3.12** [0.50-19.38] | **1.15** [0.50-3.21] | 4.78 [0.50-22.94] | 1.20 [0.50-5.90] | **1.13** [0.50-3.21] | 4.78 [0.50-22.94] |

\* Manual cleaning was not required for the fine-tuned nnU-Net model due to its high specificity, and therefore its results are not included in this column. Bolded values indicate the best performance within each evaluation stage.

Qualitative evaluation further illustrates the segmentation performance of the three models (Figure 2). In the well-segmented case (Figure 2A), all three segmentation methods produced vessel geometries closely resembling the manual reference. In the more challenging case (Figure 2B), however, DenseNet U-Net exhibited pronounced under-segmentation in the whole CoW region, while fine-tuned nnU-Net better preserved small vessel structures compared with U-Net.

**Figure 2:**
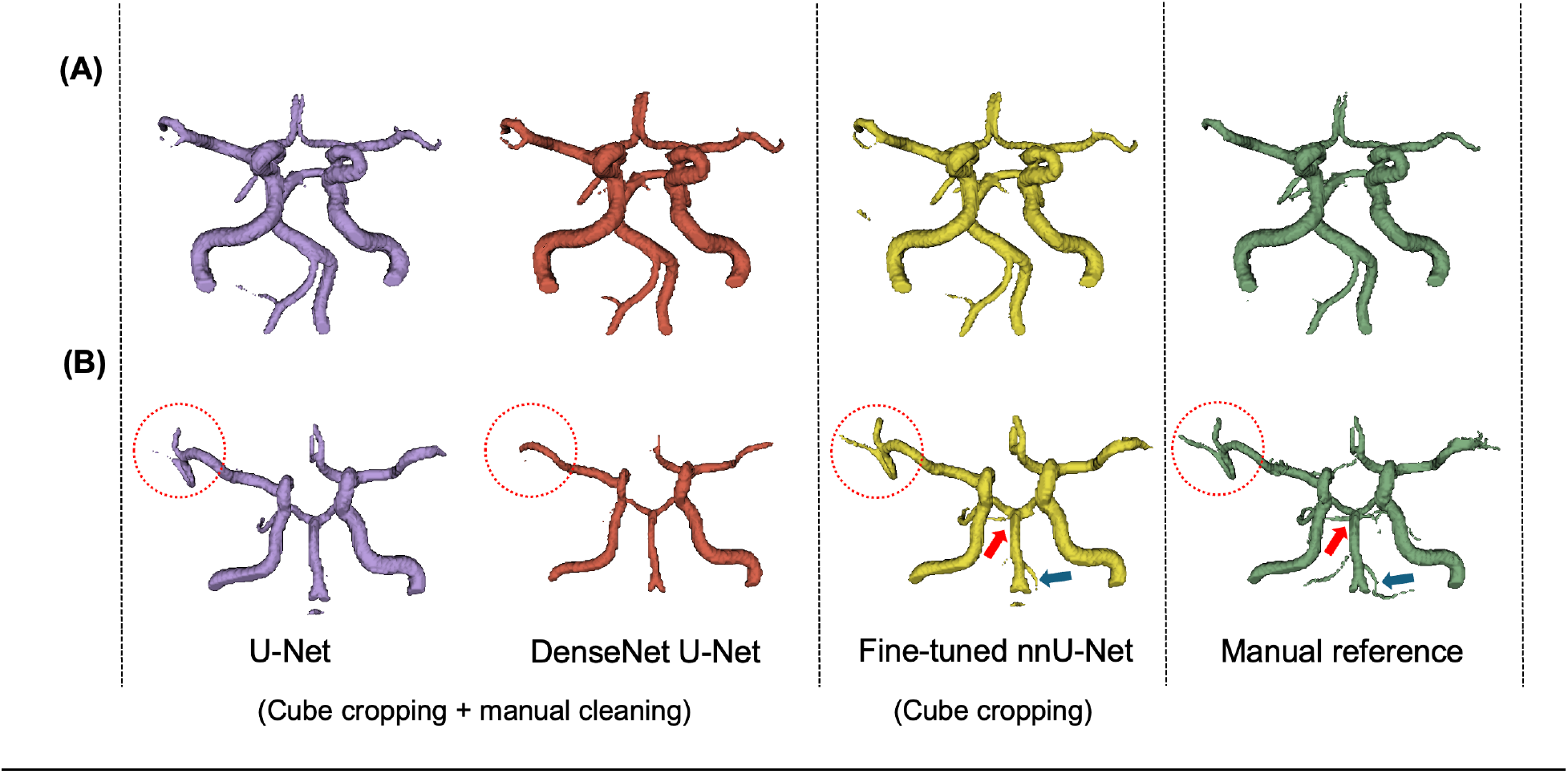
Representative 3D segmentation of the Circle of Willis (CoW) by U-Net (purple), DenseNet U-Net (red), fine-tuned nnU-Net (yellow) and manual reference (green). Panel A shows a well-segmented case with high agreement across methods (U-Net: DS = 0.893, HD95 = 0.66 mm; DenseNet U-Net: DS = 0.919, HD95 = 0.66 mm; Fine-tuned nnU-Net: DS = 0.949, HD95 = 0.66 mm). Panel B shows a more challenging case with lower agreement across methods (U-Net: DS = 0.825, HD95 = 1.00 mm; DenseNet U-Net: DS = 0.732, HD95 = 8.63 mm; Fine-tuned nnU-Net: DS = 0.852, HD95 = 0.75 mm). Red circles highlight distal vessel regions that were better preserved by the fine-tuned nnU-Net than by U-Net and DenseNet U-Net. Red and blue arrows indicate branches corresponding to the superior cerebellar artery and anterior inferior cerebellar artery, respectively, which were preserved in the fine-tuned nnU-Net segmentation and manual reference but missed by U-Net and DenseNet U-Net.

#### 3.2.2 Inter-observer agreement

For the subset selected for inter-observer analysis, Dice scores of 0.831 and 0.850 were obtained for the low- and high-resolution datasets, respectively, with corresponding HD95 values of 2.59 mm and 2.31 mm. In this same subset, non-edited model outputs were compared with the manual reference before cube cropping or manual cleaning. Compared with the inter-observer agreement, the fine-tuned nnU-Net achieved similar Dice scores (0.853 and 0.865 for the low- and high-resolution datasets, respectively) and comparable HD95 values. In contrast, DenseNet U-Net and U-Net showed lower agreement with the manual reference, particularly on the high-resolution dataset. (Table 3)

**Table 3.**
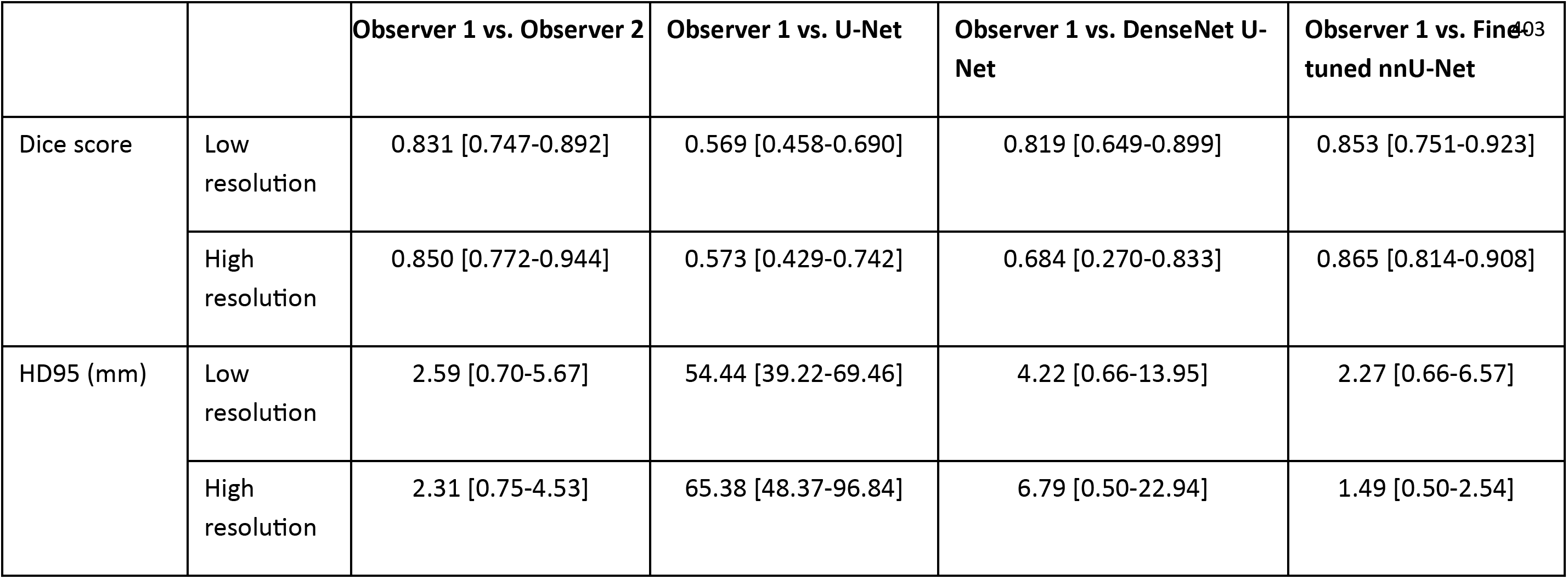
Inter-observer agreement and model performance on the subset used for inter-observer analysis. Automated model performance was evaluated using non-edited segmentation masks. All values are presented as mean [range].

#### 3.2.3 nnU-Net ablation analysis

For the nnU-Net ablation experiments, the direct-transfer nnU-Net showed substantially lower performance. When trained with all 11 cases, the from-scratch nnU-Net achieved comparable performance on the low-resolution data to the fine-tuned nnU-Net. However, on the high-resolution data, the fine-tuned nnU-Net showed slightly higher Dice and lower HD95. When comparing the fine-tuned nnU-Net with from-scratch nnU-Net models trained for 200 and 1000 epochs, a stronger performance drop was observed for both from-scratch settings when fewer than 5 training cases were used. In contrast, the fine-tuned nnU-Net showed more stable performance across reduced training sizes. Training the from-scratch nnU-Net for 1000 epochs did not consistently improve performance compared with 200 epochs, particularly on the high-resolution test set. (Supplementary Table 2 and Supplementary Figure 2)

#### 3.2.4 Study-specific subgroup analysis

Because the DenseNet U-Net, fine-tuned nnU-Net, and from-scratch nnU-Net were all trained or fine-tuned using the same 11-case low-resolution Flow@Aneurysm 4D Flow MRI dataset, we additionally evaluated their segmentation performance separately on the Flow@Aneurysm and DETAILING subsets to compare model performance across the two studies. For this subgroup analysis, the from-scratch nnU-Net refers to the model trained from scratch for 1000 epochs using the same fine-tuning set as the fine-tuned nnU-Net. The fine-tuned nnU-Net maintained consistently high Dice scores and low HD95 across both study-specific subsets. In contrast, the DenseNet U-Net showed lower performance on the DETAILING subset, particularly for the high-resolution scans. The from-scratch nnU-Net achieved comparable performance to the fine-tuned nnU-Net on the Flow@Aneurysm subset, but showed slightly lower Dice and higher HD95 on the DETAILING high-resolution subset. Overall, the fine-tuned nnU-Net showed the most stable segmentation performance across the two study-specific subsets. (Supplementary Table 3)

### 3.3 Hemodynamic analysis

First, we evaluate the flow-related metrics, including cross-sectional area, mean blood flow, and mean velocity, using their distributions (mean and SD), ICC, Bland-Altman analyses and the statistical comparison. Second, we assess the mean and maximum WSS following the same reporting structure as well as a qualitative WSS visualization.

#### 3.3.1 Flow-related metrics

*Cross-sectional area* Across all arterial segments, fine-tuned nnU-Net demonstrated the highest overall agreement with the manual reference. Its mean values were consistently closest to the manual reference across most vessels. This strong agreement was further supported by the highest ICCs in seven of nine vessels (range 0.19–0.87; exceptions: MCA-R and PCA-L). DenseNet U-Net showed the largest discrepancies from the manual reference, with ICCs (0.11–0.79) that were (near) lowest for all vessels except MCA-R and PCA-L, and a systematic underestimation of cross-sectional area compared to the manual reference with mostly negative bias and smaller means of cross-sectional area (e.g., 29.1% lower in ACA-R and 32.5% lower in ACA-L compared with the manual reference). U-Net showed an overestimation of cross-sectional area with mostly positive biases and larger means of cross-sectional areas relative to the manual reference. Notably, agreement declined in the PCAs for all methods, with the smallest ICCs observed in PCA-R/PCA-L (U-Net 0.30/0.29; DenseNet U-Net 0.31/0.11; Fine-tuned nnU-Net 0.40/0.19). (Supplementary Table 4)

*Mean blood flow* Both fine-tuned nnU-Net and U-Net showed high agreement with the manual reference, with consistently strong ICCs across vessels; Fine-tuned nnU-Net up to 0.98 in ICA-R and BA and U-Net up to 0.98 in ACA-R, ICA-R, and BA. In contrast, DenseNet U-Net yielded lower ICCs (range 0.54–0.96), particularly in PCA-R/L (0.55/0.54), and exhibited consistently negative biases, indicating systematic underestimation of blood flow relative to the manual reference. Consistent with the area analysis, all methods showed reduced ICCs in the PCA territories (U-Net 0.72/0.82; DenseNet U-Net 0.55/0.54; Fine-tuned nnU-Net 0.78/0.80). (Supplementary Table 5)

*Mean velocity* All three methods showed good agreement with the manual reference. However, unlike area and flow, which were consistently underestimated by DenseNet U-Net and overestimated by U-Net, the mean velocity showed the reverse trend with DenseNet U-Net overestimating and U-Net underestimating. DenseNet U-Net achieved the highest ICCs overall (range 0.80–0.97; e.g., MCA-R 0.97, PCA-R 0.94), with small positive biases indicating a mild overestimation of velocity (e.g., ACA-L +9.5%, BA +3.6%). Fine-tuned nnU-Net yielded comparable ICCs (0.58–0.98; e.g., ICA-R 0.97, BA 0.98) and the smallest absolute biases in most territories (typically within ±5%), making its mean velocities closest to the manual reference. U-Net showed moderate agreement (ICC 0.48–0.97) and a systematic underestimation reflected by mostly negative biases (e.g., MCA-R −10.3%, BA −9.4%). (Supplementary Table 6)

Statistical comparison revealed that the majority of vessels exhibited no significant differences (*p* > 0.05) in mean values across the three models and the manual reference for cross-sectional area, mean blood flow, and mean velocity. However, significant differences were observed in certain vessel segments. For the ACA-L, the DenseNet U-Net model showed a significant difference from the manual reference across all three parameters: cross-sectional area (*p* = 0.0339), mean blood flow (*p* = 0.0423), and mean velocity (*p* = 0.0272). For the BA, the U-Net model showed a significant difference from the other three methods for both cross-sectional area ( *p* = 0.0009 vs. DenseNet U-Net, *p* = 0.0013 vs. fine-tuned nnU-Net, and *p* = 0.0030 vs. manual reference) and mean blood flow (*p* = 0.0371 vs. DenseNet U-Net, *p* = 0.0017 vs. fine-tuned nnU-Net, and *p* = 0.0391 vs. manual reference). For mean velocity in the BA, the U-Net model also showed a significant difference from the fine-tuned nnU-Net (*p* = 0.0157) and the manual reference (*p* = 0.0096). (Figure 3)

**Figure 3:**
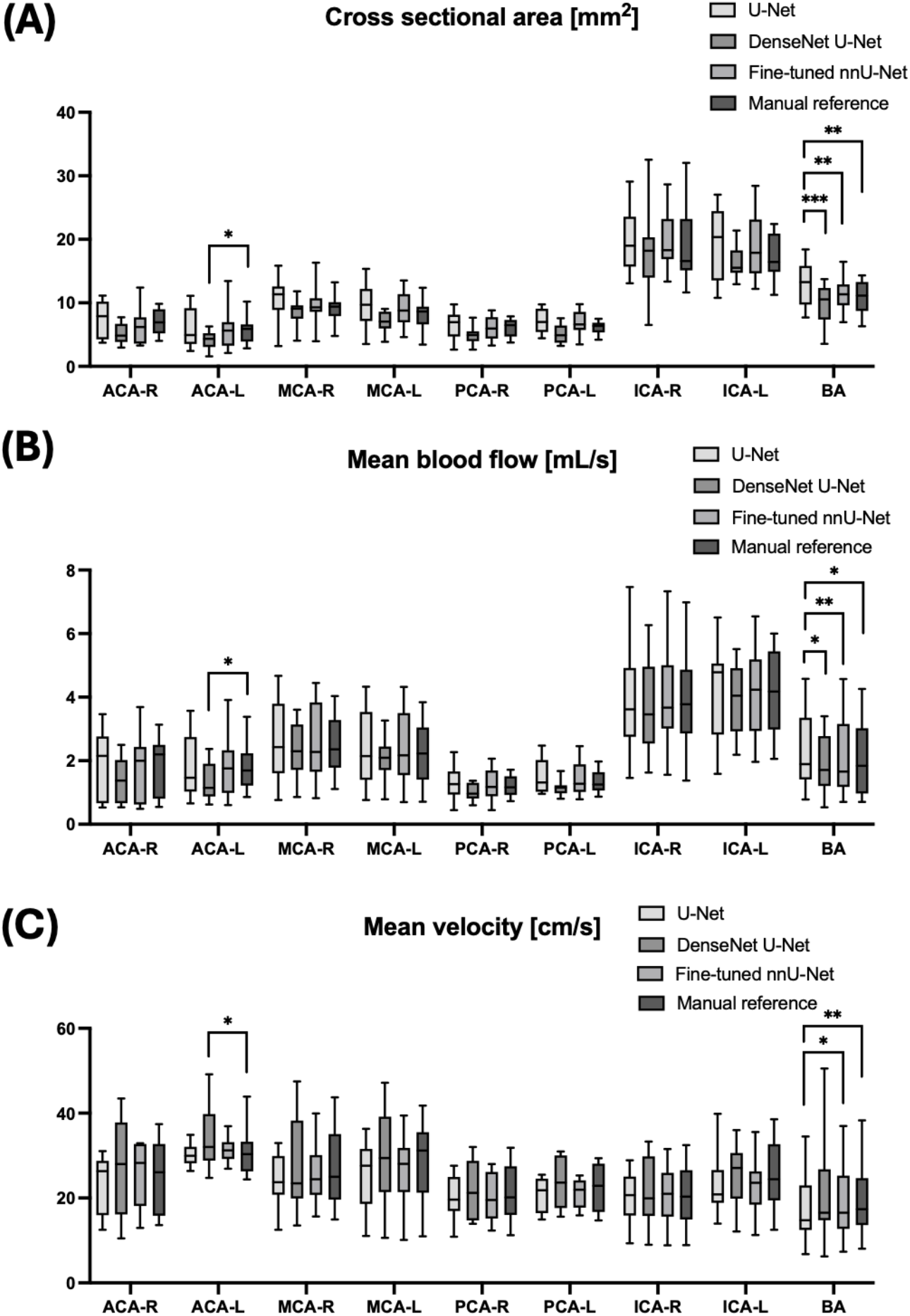
Box plots of (A) cross-sectional area (mm^2^), (B) mean blood flow (mL/s), and (C) mean velocity (cm/s) for three segmentation models and the manual reference across nine artery segments. Asterisks denote statistically significant pairwise differences (**\*** p < 0.05, **\*\***p < 0.01, **\*\*\***p < 0.001); comparisons without asterisks are not significant.

#### 3.3.2 Wall shear stress quantification

WSS quantification on segmentations from fine-tuned nnU-Net demonstrated the highest overall agreement with those computed from manual reference segmentations. Its mean WSS (1.57 ± 0.63) and maximum WSS (2.16 ± 1.05) were the closest to the manual reference (1.53 ± 0.59 and 2.09 ± 0.98), accompanied by the highest ICC values (0.96 for mean WSS, 0.97 for max WSS) and the smallest absolute biases (mean WSS:1.69%, max WSS:1.35%) among the three models (Supplementary Table 7). The Bland-Altman plots for fine-tuned nnU-Net (Figure 4C and 4F) visually confirm this, showing the tightest clustering of data points near zero.

**Figure 4:**
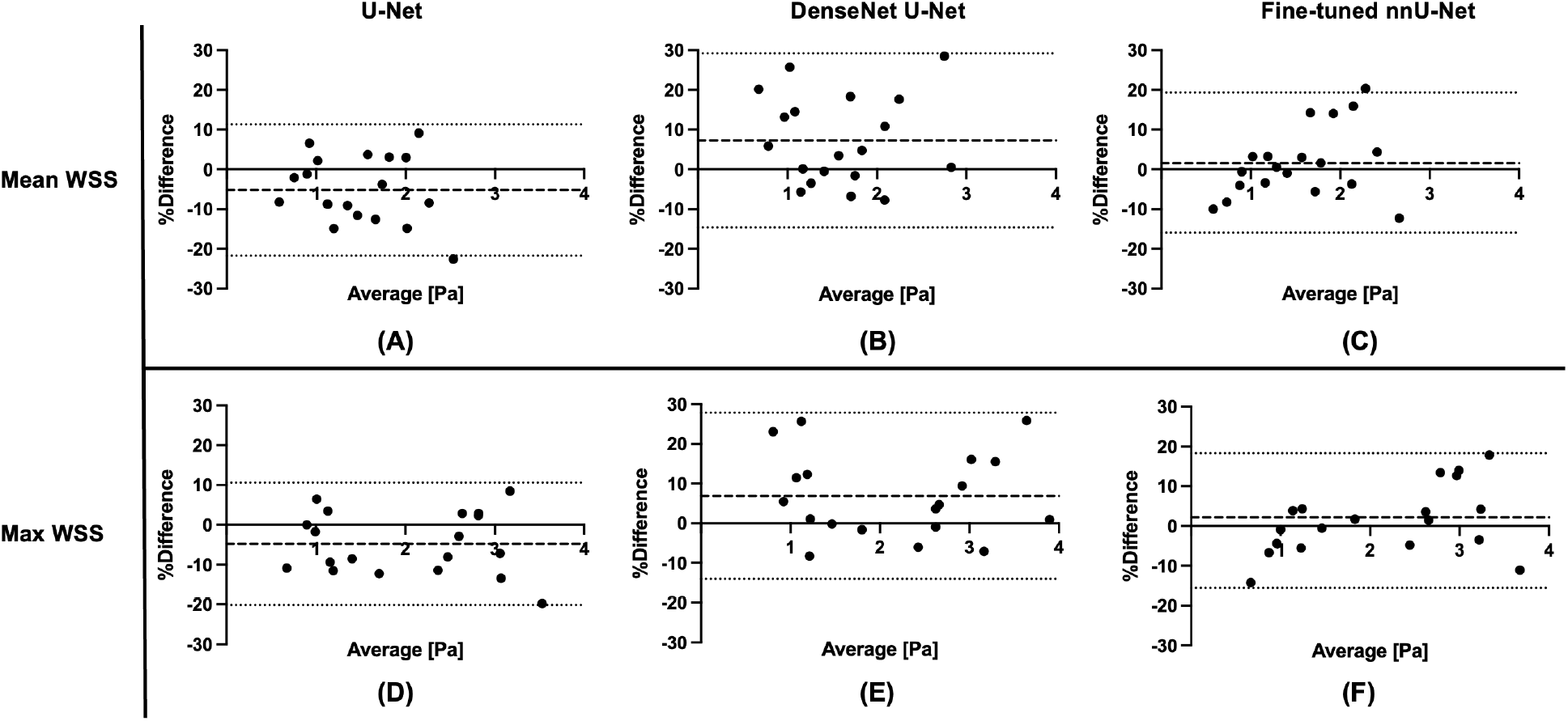
Bland-Altman plots showing the agreement of wall shear stress (WSS) measurements between three segmentation models and the manual reference. The plots illustrate the percentage difference of (A-C) mean WSS and (D-F) max WSS against the average of the two measurements. The dashed lines represent the 95% limits of agreement (LOA), and the short-dashed lines denote the mean bias.

The U-Net model showed the next highest level of agreement, with high ICC values (mean WSS: 0.95, max WSS: 0.97). However, the U-Net model exhibited a systematic negative bias, consistently underestimating WSS relative to the manual reference, with biases of −5.21% and −4.76% for mean and max WSS, respectively (Supplementary Table 7). The Bland-Altman plots (Figure 4A and 4D) visually confirm this underestimation, as the mean difference line is clearly shifted below zero.

In contrast, DenseNet U-Net had the lowest ICCs (mean WSS: 0.93, max WSS: 0.96) and a positive bias, indicating a consistent overestimation of WSS (Supplementary Table 7). The corresponding Bland-Altman plots (Figure 4B and 4E) exhibit the widest scatter and broadest LOA, reflecting the lowest consistency and the highest bias among the three models.

There were no significant differences in either mean or maximum WSS between any of the three models and the manual reference. Among them, fine-tuned nnU-Net showed the closest agreement with the manual reference, with *p* > 0.9999 for both mean and maximum WSS. In contrast, significant differences did appear in the pairwise comparisons between the three models themselves. Specifically, U-Net produced lower median mean and maximum WSS values than the other two methods, with significant differences relative to DenseNet U-Net (mean WSS: *p* = 0.0061; max WSS: *p* = 0.0059) and the fine-tuned nnU-Net (both mean and max WSS: (*p* = 0.0008). (Figure 5)

**Figure 5:**
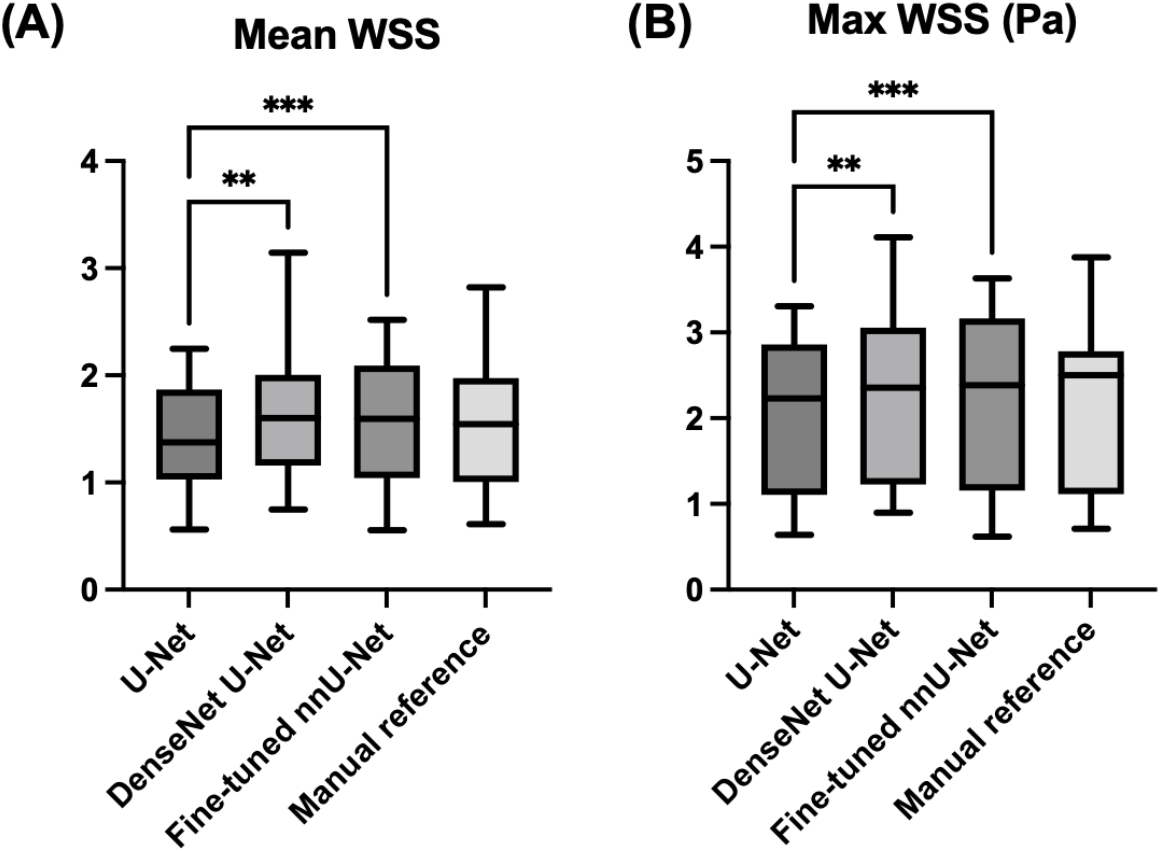
Box plots of (A) temporal mean wall shear stress (WSS, Pa) and (B) temporal max WSS, calculated from spatially averaged WSS over the entire Circle of Willis (CoW) surface, for the three segmentation models and the manual reference. Asterisks denote statistically significant pairwise differences (**\*** p < 0.05, **\*\***p < 0.01, **\*\*\***p < 0.001); comparisons without asterisks are not significant.

In addition to the quantitative metrics, WSS visualizations for a representative low-resolution case at the systolic cardiac phase were qualitatively compared across all three segmentation models and the manual reference (Figure 6). All three models successfully reproduced the expected physiological WSS patterns, with lower values observed in the larger, proximal vessels (e.g., ICA and BA) and higher values in the smaller, more distal arteries (e.g., PCA and ACA).

**Figure 6:**
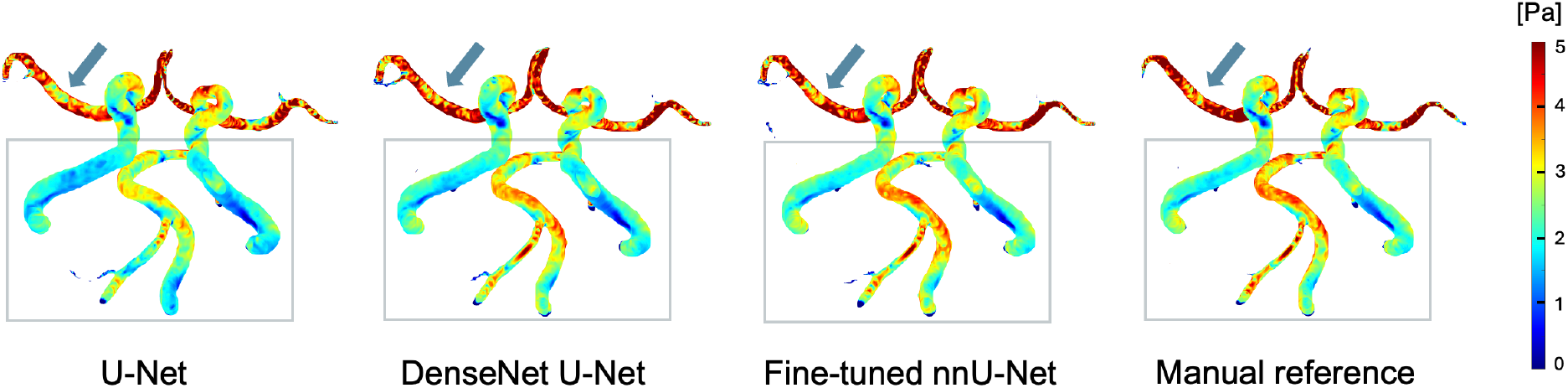
Qualitative assessment of wall shear stress (WSS) visualizations from a representative low-resolution case at the peak-systolic phase. The figure compares the spatial distribution of WSS on the 3D segmentation masks from U-Net, DenseNet U-Net, fine-tuned nnU-Net, and the manual reference. The color bar indicates WSS magnitude (Pa), with blue representing low values and red representing high values. Notably, within the regions highlighted with boxes and arrows, the spatial distribution and magnitude of WSS in the fine-tuned nnU-Net and DenseNet U-Net closely match the manual reference, whereas U-Net shows a generally cooler and smoother appearance, suggesting a slight underestimation of WSS.

## 4 Discussion

### 4.1 Principal findings

This study demonstrated that the 3D full-resolution fine-tuned nnU-Net model, pretrained on a large-scale TOF-MRA dataset and fine-tuned on a small 4D Flow MRI cohort, achieved the most robust performance in segmenting the CoW in 4D Flow MRI and in the subsequent hemodynamic quantifications. Without any manual intervention, the fine-tuned nnU-Net outperformed the previously U-Net and DenseNet U-Net in terms of segmentation accuracy. Notably, even though the fine-tuned nnU-Net used only low-resolution 4D Flow MRI data for fine-tuning, it achieved comparable segmentation performance on the high-resolution data, indicating strong generalization across different spatial resolutions and acquisition protocols. Furthermore, our results revealed that segmentation accuracy directly influenced the precision of hemodynamic quantification. Among the three segmentation methods, the fine-tuned nnU-Net demonstrated the highest agreement with the manual reference, while errors in vessel delineation from U-Net and DenseNet U-Net propagated to systematic biases in blood flow and WSS estimation. These findings highlight the importance of reliable vessel segmentation for accurate 4D Flow MRI–based hemodynamic analysis.

### 4.2 Segmentation performance and influencing factors

Until recently, most work on intracranial artery segmentation has focused on imaging modalities with higher spatial resolution and signal quality, such as TOF-MRA, CTA, and DSA (Kim et al., 2025; Min et al., 2025; Su et al., 2024). In contrast, 4D Flow MRI typically has lower spatial resolution, and its relatively long acquisition time increases the risk of motion artifacts, further degrading image quality. Consequently, segmentation research in 4D Flow MRI has concentrated on large vessels such as the aorta and pulmonary arteries, where partial-volume effects are less severe relative to lumen size. Several deep learning approaches have been developed in that setting, including the DenseNet U-Net, which was originally designed for aortic segmentation (Aviles et al., 2021; Berhane et al., 2020; Manokaran et al., 2023). Winter et al. later applied DenseNet U-Net to intracranial vessel segmentation and achieved a DS around 0.85 for the CoW when trained on over 100 4D Flow MRI scans (Winter et al., 2024). However, because the trained model weights were not publicly available, we trained the DenseNet U-Net from scratch on our own 4D Flow MRI data. While training the same architecture from scratch on our limited 7T cohort (11 patients) did not achieve comparable performance, the fine-tuned nnU-Net achieved a DS around 0.86 using the same cohort for fine-tuning. Additionally, extending DenseNet U-Net training from 100 to 300 epochs as in Winter et al. did not consistently improve segmentation performance (supplementary table 8) (Winter et al., 2024). In contrast, the pretrained U-Net baseline was applied as a direct-transfer model from TOF-MRA to 4D Flow MRI without domain-specific fine-tuning. Therefore, the performance difference between U-Net and the fine-tuned nnU-Net may be largely influenced by the lack of domain adaptation in addition to differences in the model framework.

The nnU-Net ablation experiments further clarified the role of TOF-MRA pre-training and 4D Flow MRI fine-tuning. Although both modalities are flow-sensitive, TOF-MRA primarily relies on inflow-related enhancement, whereas 4D Flow MRI-derived PCMRA combines magnitude and velocity information, potentially highlighting additional vascular structures, including veins and extracranial vessels. Furthermore, TOF-MRA retains some anatomical background information from stationary brain tissue despite RF saturation, while PCMRA largely suppresses stationary tissue signals, resulting in less visible anatomical background structures. These differences in both vascular appearance and surrounding tissue contrast may affect the transferability of learned features between the two modalities. Indeed, visual inspection of the direct-transfer nnU-Net predictions suggested that some segmentation errors arose from false-positive predictions of non-target vascular structures rather than inaccurate delineation of intracranial arteries alone. These observations may partly explain why TOF-MRA pre-training alone was insufficient for direct application to 4D Flow MRI and why domain-specific fine-tuning improved segmentation performance. With all 11 training cases, the from-scratch nnU-Net achieved slightly better mean Dice and HD95 on the low-resolution test set, whereas the fine-tuned nnU-Net performed better on the high-resolution test set. Since fine-tuning was performed exclusively on low-resolution 4D Flow MRI, this suggests that TOF-MRA pre-training may have improved generalization to acquisition settings not represented in the fine-tuning data. However, this benefit cannot be attributed to spatial resolution alone, as the low- and high-resolution datasets also differed in VENC and other acquisition parameters. The fine-tuned nnU-Net also showed more stable performance on the high-resolution test set when fewer training cases were available. In addition, extending from-scratch training from 200 to 1000 epochs did not consistently improve performance, particularly on the high-resolution test set, suggesting that longer training alone may not improve robustness in limited-data settings. Together, these findings suggest that TOF-MRA pre-training followed by 4D Flow MRI fine-tuning may help achieve robust segmentation performance with limited 4D Flow MRI annotations. Because these ablation experiments used a single nested subset sequence without repeated random sampling, the observed robustness across reduced fine-tuning subsets should be interpreted as exploratory. In addition to this robustness, the study-specific subgroup analysis suggests potential generalizability across acquisition protocols, as the fine-tuned nnU-Net showed more stable performance across the Flow@Aneurysm and DETAILING subsets than the DenseNet U-Net and from-scratch nnU-Net. However, this observation should be interpreted cautiously given the small size of the DETAILING subset and should be validated in larger cohorts.

When interpreting the segmentation performance, several factors beyond model architecture and training strategy should be considered. First, differences in acquisition may have influenced the input image characteristics, including different VENC values within our dataset, as well as differences in VENC strategy and field strength when comparing our study with Winter et al. Within our dataset, different VENC values were used across the low- and high-resolution acquisitions. The high-resolution datasets were acquired with a lower VENC (50 cm/s), which may improve the characterisation of slow-flow components in PCMRA images, but also increases susceptibility to velocity aliasing in faster-flow regions and may contribute to locally heterogeneous vessel signal. Therefore, differences between the low- and high-resolution datasets may reflect not only spatial resolution but also VENC-related differences in PCMRA appearance, which may in turn influence segmentation performance. In contrast to Winter et al., who used dual-VENC acquisitions at 3T, our study used single-VENC acquisitions at 7T. Dual-VENC imaging provides the benefits of a broader velocity dynamic range and higher SNR, particularly when segmenting both arterial and venous structures with a wide range of flow velocities (Aviles et al., 2021). However, our study only focused on the arteries within the CoW, which may reduce the relative importance of the broader velocity coverage provided by dual-VENC imaging.

Compared with 3T, 7T MRI is more susceptible to B1 field inhomogeneity, which may result in spatially variable flip angles, signal amplitudes, and local SNR (Schmitter et al., 2016). No dedicated B1 correction was applied in the present study. Therefore, B1 field inhomogeneity at 7T may have contributed to spatial variations in signal intensity in the PCMRA images. In addition, complex flow patterns, particularly in or near aneurysmal regions, may cause intravoxel dephasing and flow-related signal dropouts, potentially reducing local vessel conspicuity in PCMRA images. However, the higher field strength also provides increased SNR, which can be used to acquire images with higher spatial resolution (Gottwald, Töger, et al., 2020). Accordingly, our data included near-isotropic 0.7 mm and isotropic 0.5 mm acquisitions, in contrast to the approximately 1 mm spatial resolution used in Winter et al. (Winter et al., 2024). Second, the preprocessing pipelines differed across the three evaluated models, including differences in intensity normalization, patch-based processing, fixed cropping or padding, and sliding-window inference. Each model was evaluated using the preprocessing pipeline corresponding to its published or default implementation to preserve methodological consistency as much as possible. Nevertheless, these implementation differences may have contributed to the observed performance differences in addition to the network architectures themselves. An additional consideration when interpreting the segmentation results is the uncertainty associated with the manual reference. Inter-observer analysis demonstrated that manual segmentations themselves exhibit variability, indicating that the reference standard is not perfectly reproducible. Therefore, part of the observed difference may arise from uncertainty in the reference segmentation itself.

### 4.3 Impact of segmentation and methodological factors on hemodynamic quantification

The bias pattern in vessel area closely aligns with the errors observed in flow and velocity. DenseNet U-Net consistently underestimated cross-sectional areas across multiple vessels, showing negative bias and the lowest ICC values for this metric. Consequently, its mean flow values were also negatively biased. In contrast, U-Net tended to overestimate lumen size, producing positive area bias and systematically underestimating velocity and WSS. Although DenseNet U-Net showed the best agreement with the manual reference for mean velocity, as reflected by generally higher ICC, this does not necessarily indicate superior performance. This apparent agreement largely resulted from its underestimation of vessel area and corresponding flow, which brought the computed mean velocity closer to the manual reference. By comparison, the fine-tuned nnU-Net exhibited smaller and more balanced area biases, leading to the highest overall agreement with the manual reference for both flow and WSS.

Notably, all models showed relatively low ICC values for the cross-sectional area and mean blood flow of the PCAs. This can be explained by the principle of PCMRA images, which combines magnitude and velocity information to generate flow-dependent contrast. Because signal intensity in PCMRA is proportional to local flow velocity, vessels with slower flow appear darker and less contrasted. Given that the PCA is relatively slow-flowing, the flow contrast on PCMRA images is reduced, which increases segmentation difficulty. Even minor radius deviations in such small vessels can cause large relative changes in cross-sectional area and consequently affect flow measurement.

Physiological variability, such as sex-, age-, and side-related differences, may also affect cerebral hemodynamics (Bakker et al., 2004; Marinoni et al., 1998; Wu et al., 2017). For instance, Fico et al. reported a mean blood flow difference of approximately 0.4 mL/s between young and older adults in the left M1 segment of MCA measured by 4D Flow MRI (Fico et al., 2023). Notably, the mean blood flow derived from different segmentation models in our study showed a similar magnitude of variation (e.g., U-Net 2.45 mL/s versus DenseNet U-Net 2.02 mL/s in the left MCA; Δ = 0.43 mL/s). This indicates that the variability in hemodynamics introduced by the different segmentation models may equal or exceed the biological effects, underscoring the importance of precise vessel delineation for reliable hemodynamic quantification.

A further contributor to hemodynamic differences is the fact that dynamic flow fields are evaluated on a static segmentation. Blood flow is pulsatile, and intracranial arteries are compliant, changing their lumen throughout the cardiac cycle, whereas the manual reference used in our analysis was derived from a time-averaged PCMRA image that cannot capture phase-specific vessel geometry. Although the segmentation models could in principle generate time-resolved masks, producing a time-resolved manual reference would require frame-by-frame manual annotation and extensive quality control, which is impractical. In addition, because centerlines and analysis planes were generated separately for each segmentation, the observed hemodynamic differences may reflect both vessel segmentation differences and variability introduced by plane placement. However, this approach reflects a realistic downstream workflow, and variability was minimized as much as possible by selecting analysis planes at corresponding anatomical locations across methods. In addition, QVT-based flow analysis was restricted to vessel segments that could be successfully analyzed across all four segmentation pipelines, the reported agreement may be overestimated and does not account for segmentation or analysis failures. Furthermore, partial-volume effects and flow-related signal loss cause uncertainty in both the vessel boundary and the near-wall velocity gradient, thereby affecting not only segmentation accuracy but also WSS estimation. Therefore, both manual and automated PCMRA-based segmentations can deviate from the physical vessel wall, and the reported WSS agreement should be interpreted as agreement relative to the reference segmentation.

### 4.4 Limitations and future work

Our study has some limitations that also highlight paths for future research. First, since the orientation and position of scan volumes were adjusted according to the aneurysm locations, there were minor variations in the coverage of CoW. Combined with the relatively small 4D Flow MRI dataset, this may have limited the model‘s ability to learn robust and generalizable representations. Although transfer learning from a large public TOF-MRA dataset may have mitigated this issue to some extent, future work should evaluate this effect on a larger and more diverse 4D Flow MRI cohort.

Second, the comparison with baseline models should be interpreted as a pipeline-level comparison rather than a fully architecture-matched comparison. This design was motivated by the available implementations of the baseline models and by our aim to preserve their published or default settings as much as possible. Therefore, the evaluated baseline models differed not only in architecture but also in training strategy and preprocessing pipeline. Future studies should include architecture-matched transfer-learning experiments to further isolate the effects of model architecture and training strategy.

Third, future studies could include topology-aware metrics, such as clDice, or centerline-based measures to better evaluate vessel connectivity and small-branch preservation in branching vascular trees. Furthermore, this framework was developed and evaluated using 7T data, whereas most clinical 4D Flow MRI acquisitions are performed at 3T and often at lower spatial resolution. Therefore, future work should evaluate whether the transfer-learning strategy can be adapted to 3T 4D Flow MRI through dedicated fine-tuning on representative clinical datasets.

Finally, although most subjects in the 4D Flow MRI cohorts were patients with IAs, our segmentation focused only on the intracranial arteries and did not include the aneurysm sacs. Previous work has demonstrated the feasibility of automated aneurysm segmentation and WSS quantification using 4D Flow MRI (Winter et al., 2025). However, obtaining reliable manual reference segmentations of aneurysm sacs was challenging in our dataset, particularly for smaller aneurysms with heterogeneous signal and pronounced flow-related signal loss, limiting the reliable evaluation of aneurysm-specific segmentation accuracy and WSS measurements. Given the potential clinical value of IA segmentation and aneurysm-specific hemodynamic assessment for treatment planning and risk evaluation, extending the current framework to jointly segment intracranial arteries and aneurysm sacs represents an important direction for future research.

## 5 Conclusion

In this study, we demonstrated that a 3D full-resolution nnU-Net model pretrained on TOF-MRA and fine-tuned on 7T 4D Flow MRI data, provides a robust and accurate solution for automated intracranial artery segmentation and hemodynamic quantification from 4D Flow MRI. The fine-tuned nnU-Net achieved accurate CoW segmentation without manual editing and provides a promising framework for 4D Flow MRI–based cerebrovascular analysis, supporting future clinical and research applications. However, because reference-informed ROI restriction was used in the present validation framework to standardize downstream comparisons across segmentation pipelines, the current workflow with ROI cube cropping should be interpreted as an evaluation framework rather than a fully automated clinical pipeline. Future work will focus on validating our approach on larger, multi-center 4D Flow MRI cohorts, exploring aligned training strategies across different architectures, including clinical 3T datasets, and extending the framework to aneurysm segmentation and analysis.

## Data Availability

Due to ethical and institutional privacy restrictions, the clinical imaging data used in this study cannot be shared publicly. The trained model weights and configuration files necessary for inference will be released on GitHub upon publication.

## Acknowledgement

AI-assisted tools (ChatGPT, OpenAI) were used solely to support language editing and phrasing. These tools did not generate scientific content, data, interpretations, or conclusions. All scientific content, interpretations, results, and final wording were produced by the authors and reviewed and approved by all authors.

## Author Contributions

JZ, KMN, ICvdS, and CMWT conceived the study and designed the research framework. JZ conducted the study, performed the data analysis, and drafted the initial manuscript. ICvdS and CMWT supervised the work and acquired funding for the project. ASV contributed to the methodological development. KMN participated in data acquisition and reconstruction, and MB assisted with data collection. EMS and PvO provided analytical software and technical guidance. All authors contributed to the revision of the manuscript and approved the final version.

## Reference

Andre, J. B., Bresnahan, B. W., Mossa-Basha, M., Hoff, M. N., Smith, C. P., Anzai, Y., & Cohen, W. A. (2015). Toward Quantifying the Prevalence, Severity, and Cost Associated With Patient Motion During Clinical MR Examinations. Journal of the American College of Radiology, 12(7), 689–695. 10.1016/j.jacr.2015.03.007

Aviles, J., Talou, G. D. M., Camara, O., Córdova, M. M., Ferez, X. M., Romero, D., Ferdian, E., Gilbert, K., Elsayed, A., Young, A. A., Dux-Santoy, L., Ruiz-Munoz, A., Teixido-Tura, G., Rodriguez-Palomares, J., & Guala, A. (2021). Domain Adaptation for Automatic Aorta Segmentation of 4D Flow Magnetic Resonance Imaging Data from Multiple Vendor Scanners. In D. B. Ennis, L. E. Perotti, & V. Y. Wang (Eds.), Functional Imaging and Modeling of the Heart (pp. 112–121). Springer International Publishing. 10.1007/978-3-030-78710-3_12

Bakker, S. L. M., de Leeuw, F.-E., den Heijer, T., Koudstaal, P. J., Hofman, A., & Breteler, M. M. B. (2004). Cerebral Haemodynamics in the Elderly: The Rotterdam Study. Neuroepidemiology, 23(4), 178–184. 10.1159/000078503

Berhane, H., Scott, M., Elbaz, M., Jarvis, K., McCarthy, P., Carr, J., Malaisrie, C., Avery, R., Barker, A. J., Robinson, J. D., Rigsby, C. K., & Markl, M. (2020). Fully automated 3D aortic segmentation of 4D flow MRI for hemodynamic analysis using deep learning. Magnetic Resonance in Medicine, 84(4), 2204–2218. 10.1002/mrm.28257

Busch, J., Giese, D., & Kozerke, S. (2017). Image-based background phase error correction in 4D flow MRI revisited. Journal of Magnetic Resonance Imaging, 46(5), 1516–1525. 10.1002/jmri.25668

de Vos, V., Timmins, K. M., van der Schaaf, I. C., Ruigrok, Y., Velthuis, B. K., & Kuijf, H. J. (2021). Automatic cerebral vessel extraction in TOF-MRA using deep learning. Medical Imaging 2021: Image Processing, 11596, 651–660.

Dunås, T., Holmgren, M., Wåhlin, A., Malm, J., & Eklund, A. (2019). Accuracy of blood flow assessment in cerebral arteries with 4D flow MRI: Evaluation with three segmentation methods. Journal of Magnetic Resonance Imaging: JMRI, 50(2), 511–518. 10.1002/jmri.26641

Fico, B. G., Miller, K. B., Rivera-Rivera, L. A., Corkery, A. T., Pearson, A. G., Loggie, N. A., Howery, A. J., Rowley, H. A., Johnson, K. M., Johnson, S. C., Wieben, O., & Barnes, J. N. (2023). Cerebral hemodynamics comparison using transcranial doppler ultrasound and 4D flow MRI. Frontiers in Physiology, 14. 10.3389/fphys.2023.1198615

Gottwald, L. M., Peper, E. S., Zhang, Q., Coolen, B. F., Strijkers, G. J., Nederveen, A. J., & van Ooij, P. (2020). Pseudo-spiral sampling and compressed sensing reconstruction provides flexibility of temporal resolution in accelerated aortic 4D flow MRI: A comparison with k-t principal component analysis. NMR in Biomedicine, 33(4), e4255.

Gottwald, L. M., Töger, J., Markenroth Bloch, K., Peper, E. S., Coolen, B. F., Strijkers, G. J., van Ooij, P., & Nederveen, A. J. (2020). High Spatiotemporal Resolution 4D Flow MRI of Intracranial Aneurysms at 7T in 10 Minutes. AJNR. American Journal of Neuroradiology, 41(7), 1201–1208. 10.3174/ajnr.A6603

Hennemuth, A., Friman, O., Schumann, C., Bock, J., Drexl, J., Huellebrand, M., Markl, M., & Peitgen, H.-O. (2011). Fast interactive exploration of 4D MRI flow data. Medical Imaging 2011: Visualization, Image-Guided Procedures, and Modeling, 7964, 110–120.

Isensee, F., Jaeger, P. F., Kohl, S. A. A., Petersen, J., & Maier-Hein, K. H. (2021). nnU-Net: A self-configuring method for deep learning-based biomedical image segmentation. Nature Methods, 18(2), 203–211. 10.1038/s41592-020-01008-z

Kim, H., Seo, K.-H., Kim, K., Shim, J., & Lee, Y. (2025). Application and optimization of the U-Net++ model for cerebral artery segmentation based on computed tomographic angiography images. European Journal of Radiology, 188, 112137. 10.1016/j.ejrad.2025.112137

Köhler, B., Born, S., van Pelt, R. F., Hennemuth, A., Preim, U., & Preim, B. (2017). A survey of cardiac 4D PC-MRI data processing. Computer Graphics Forum, 36(6), 5–35.

Manokaran, J., Garcia, J., & Ukwatta, E. (2023). Automated aortic segmentation and quantification of hemodynamic parameters from 4D flow MRI using deep learning techniques. Journal of Medical Imaging, 10(06). 10.1117/1.JMI.10.6.066001

Marinoni, M., Ginanneschi, A., Inzitari, D., Mugnai, S., & Et., A. (1998). Sex-related differences in human cerebral hemodynamics. Acta Neurologica Scandinavica. 10.1111/j.1600-0404.1998.tb05961.x

Markl, M., Frydrychowicz, A., Kozerke, S., Hope, M., & Wieben, O. (2012). 4D flow MRI. Journal of Magnetic Resonance Imaging, 36(5), 1015–1036. 10.1002/jmri.23632

Min, Y., Li, J., Jia, S., Li, Y., & Nie, S. (2025). Automated Cerebrovascular Segmentation and Visualization of Intracranial Time-of-Flight Magnetic Resonance Angiography Based on Deep Learning. Journal of Imaging Informatics in Medicine, 38(2), 703–716. 10.1007/s10278-024-01215-6

Mou, L., Lin, J., Zhao, Y., Liu, Y., Ma, S., Zhang, J., Lv, W., Zhou, T., Frangi, A. F., & Zhao, Y. (2023). COSTA: A Multi-center Multi-vendor TOF-MRA Dataset and A Novel Cerebrovascular Segmentation Network. IEEE Transactions on Medical Imaging.

Roberts, G. S., Hoffman, C. A., Rivera-Rivera, L. A., Berman, S. E., Eisenmenger, L. B., & Wieben, O. (2023). Automated hemodynamic assessment for cranial 4D flow MRI. Magnetic Resonance Imaging, 97, 46–55.

Rothenberger, S. M., Patel, N. M., Zhang, J., Schnell, S., Craig, B. A., Ansari, S. A., Markl, M., Vlachos, P. P., & Rayz, V. L. (2023). Automatic 4D Flow MRI Segmentation Using the Standardized Difference of Means Velocity. IEEE Transactions on Medical Imaging, 42(8), 2360–2373. 10.1109/TMI.2023.3251734

Schmitter, S., Schnell, S., Uğurbil, K., Markl, M., & Van de Moortele, P.-F. (2016). Towards high-resolution 4D flow MRI in the human aorta using kt-GRAPPA and B1+ shimming at 7T. Journal of Magnetic Resonance Imaging, 44(2), 486–499. 10.1002/jmri.25164

Schrauben, E., Wåhlin, A., Ambarki, K., Spaak, E., Malm, J., Wieben, O., & Eklund, A. (2015). Fast 4D flow MRI intracranial segmentation and quantification in tortuous arteries. Journal of Magnetic Resonance Imaging, 42(5), 1458–1464.

Sobisch, J., Bizjak, Ž., Chien, A., & Špiclin, Ž. (2022). Automated intracranial vessel labeling with learning boosted by vessel connectivity, radii and spatial context. In E. Bekkers, J. M. Wolterink, & A. Aviles-Rivero (Eds.), Proceedings of the First International Workshop on Geometric Deep Learning in Medical Image Analysis (Vol. 194, pp. 34–44). PMLR. https://proceedings.mlr.press/v194/sobisch22a.html

Su, R., van der Sluijs, P. M., Chen, Y., Cornelissen, S., van den Broek, R., van Zwam, W. H., van der Lugt, A., Niessen, W. J., Ruijters, D., & van Walsum, T. (2024). CAVE: Cerebral artery-vein segmentation in digital subtraction angiography. Computerized Medical Imaging and Graphics: The Official Journal of the Computerized Medical Imaging Society, 115, 102392. 10.1016/j.compmedimag.2024.102392

Tuijl, R. J. van, Hertog, C. S. den, Timmins, K. M., Velthuis, B. K., Ooij, P. van, Zwanenburg, J. J. M., Ruigrok, Y. M., & Schaaf, I. C. van der. (2024). Intra-Aneurysmal High-Resolution 4D MR Flow Imaging for Hemodynamic Imaging Markers in Intracranial Aneurysm Instability. American Journal of Neuroradiology, 45(11), 1678–1684. 10.3174/ajnr.A8380

Uecker, M., Tamir, J. I., Ong, F., & Lustig, M. (2016). The BART toolbox for computational magnetic resonance imaging. Proc Intl Soc Magn Reson Med, 24, 1.

van Ooij, P., Potters, W. V., Guédon, A., Schneiders, J. J., Marquering, H. A., Majoie, C. B., VanBavel, E., & Nederveen, A. J. (2013). Wall shear stress estimated with phase contrast MRI in an in vitro and in vivo intracranial aneurysm. Journal of Magnetic Resonance Imaging, 38(4), 876–884.

van Ooij, P., Zwanenburg, J. J. M., Visser, F., Majoie, C. B., vanBavel, E., Hendrikse, J., & Nederveen, A. J. (2013). Quantification and visualization of flow in the Circle of Willis: Time-resolved three-dimensional phase contrast MRI at 7 T compared with 3 T. Magnetic Resonance in Medicine, 69(3), 868–876. 10.1002/mrm.24317

Van Tuijl, R. J., Timmins, K. M., Velthuis, B. K., Van Ooij, P., Zwanenburg, J. J. M., Ruigrok, Y. M., & Van Der Schaaf, I. C. (2024). Hemodynamic Parameters in the Parent Arteries of Unruptured Intracranial Aneurysms Depend on Aneurysm Size and Are Different Compared to Contralateral Arteries: A 7 Tesla 4D Flow MRI Study. Journal of Magnetic Resonance Imaging, 59(1), 223–230. 10.1002/jmri.28756

Winter, P., Berhane, H., Moore, J. E., Aristova, M., Reichl, T., Wollenberg, J., Richter, A., Jarvis, K. B., Patel, A., Caprio, F. Z., Abdalla, R. N., Ansari, S. A., Markl, M., & Schnell, S. (2024). Automated intracranial vessel segmentation of 4D flow MRI data in patients with atherosclerotic stenosis using a convolutional neural network. Frontiers in Radiology, 4. 10.3389/fradi.2024.1385424

Winter, P., Berhane, H., Aristova, M., Ansari, S., Markl, M., & Schnell, S. (2025, August 21–24). Automatic segmentation and wall shear stress quantification of intracranial aneurysms from 4D flow using convolutional neural networks [Conference abstract]. 37th Annual International Meeting of the Society for Magnetic Resonance Angiography, Budapest, Hungary. https://society4mra.org/download/smra-2025-abstracts/

Wu, C., Schnell, S., Vakil, P., Honarmand, A. R., Ansari, S. A., Carr, J., Markl, M., & Prabhakaran, S. (2017). In Vivo Assessment of the Impact of Regional Intracranial Atherosclerotic Lesions on Brain Arterial 3D Hemodynamics. American Journal of Neuroradiology, 38(3), 515–522. 10.3174/ajnr.A5051

