## Supplementary Material for "Automated Segmentation of Intracranial Arteries on 4D Flow MRI for Hemodynamic Quantification"

***Supplementary Table 1:*** *Detailed scan parameters of 4D Flow MRI datasets.*

| **Dataset** | **Slice orientation** | **FOV (AP×RL×FH)**  **(mm^3^)** | **Resolution**  **(AP×RL×FH)**  **(mm^3^)** | **TR/TE**  **(ms/ms)** | **Acceleration factor** | **Flip angle (⁰)** | **VENC (cm/s)** | **Reconstructed cardiac phases** |
| --- | --- | --- | --- | --- | --- | --- | --- | --- |
| Flow@Aneurysm 1 | Angulated coronal | 30 × 190 × 190 | 0.7 × 0.66 × 0.66 | 6.4/2.2 | 7 | 15° | 100 | 12 |
| Flow@Aneurysm 2 | Angulated coronal | 20 × 190 × 190 | 0.5 × 0.5 × 0.5 | 7.0/2.2 | 7 | 10° | 50 | 12 |
| DETAILING 1 | Angulated transverse | 194 × 183 × 42 | 0.75 × 0.75 × 0.75 | 5.2/1.98 | 9 | 15° | 100 | 12 |
| DETAILING 2 | Angulated transverse | 192 × 180 × 21 | 0.5 × 0.5 × 0.5 | 6.4/3.1 | 9 | 15° | 50 | 12 |

**Note:** Datasets labeled ‘1’ and ‘2’ correspond to low-resolution and high-resolution 4D Flow MRI acquisitions, respectively.

***Supplementary Table 2:*** *Segmentation performance metrics for the nnU-Net ablation models. HD95 indicates the 95% Hausdorff distance. The from-scratch nnU-Net was trained for 1000 epochs using the same 11-case 4D Flow MRI fine-tuning set as the fine-tuned nnU-Net. All values are presented as mean [range].*

|  |  | **Fine-tuned**  **nnU-Net** | **From-scratch nnU-Net** | **Direct-transfer**  **nnU-Net** |
| --- | --- | --- | --- | --- |
| Dice score | Low resolution | 0.860  [0.732-0.946] | 0.867  [0.785-0.949] | 0.681  [0.507-0.886] |
|  | High resolution | 0.856  [0.739-0.927] | 0.847  [0.733-0.918] | 0.690  [0.439-0.809] |
| HD95  (mm) | Low resolution | 2.98  [0.66-28.04] | 1.83 [0.66-12.01] | 40.00  [8.73-72.26] |
|  | High resolution | 3.12  [0.50-19.38] | 5.06  [0.50-20.69] | 53.58  [9.62-95.03] |

***Supplementary Table 3:*** *Study-specific segmentation performance of the evaluated models in the Flow@Aneurysm and DETAILING subsets. The from-scratch nnU-Net was trained for 1000 epochs using the same 11-case 4D Flow MRI fine-tuning set as the fine-tuned nnU-Net. All values are presented as mean [range].*

|  |  | DenseNet U-Net | | Fine-tuned nnU-Net | | From-scratch nnU-Net | |
| --- | --- | --- | --- | --- | --- | --- | --- |
|  |  | Flow@Aneurysm | DETAILING | Flow@Aneurysm | DETAILING | Flow@Aneurysm | DETAILING |
| Dice score | Low resolution | 0.850  [0.726-0.919] | 0.709  [0.649-0.801] | 0.870  [0.732-0.946] | 0.831  [0.751-0.876] | 0.878  [0.807-0.949] | 0.838  [0.785-0.868] |
|  | High resolution | 0.813  [0.661-0.869] | 0.460  [0.270-0.695] | 0.859  [0.768-0.927] | 0.846  [0.739-0.904] | 0.852  [0.766-0.918] | 0.830  [0.733-0.890] |
| HD95  (mm) | Low resolution | 3.11  [0.66-13.80] | 10.52  [6.26-14.07] | 3.32  [0.66-28.04] | 2.05  [0.67-6.57] | 1.94  [0.066-12.01] | 1.52  [0.75-3.81] |
|  | High resolution | 1.74  [0.59-4.77] | 15.66  [4.07-22.94] | 2.63  [0.50-19.38] | 4.74  [0.75-10.92] | 2.81  [0.495-20.69] | 12.57  [4.04-19.88] |

***Supplementary Table 4:*** *Cross-sectional area (mm^2^) by artery: mean ± SD [range], intraclass correlation (ICC [95% CI]), and Bland-Altman bias ± limits of agreement (LOA, %) for three segmentation models vs. manual reference.*

|  | U-Net | | | DenseNet U-Net | | | Fine-tuned nnU-Net | | | Manual reference |
| --- | --- | --- | --- | --- | --- | --- | --- | --- | --- | --- |
|  | Mean ± SD [range] | ICC  [95% CI] | Bias ± LOA [%] | Mean ± SD [range] | ICC  [95% CI] | Bias ± LOA [%] | Mean ± SD [range] | ICC  [95% CI] | Bias ± LOA [%] | Mean ± SD [range] |
| ACA-R | 7.3 ± 2.9  [3.7-11.2] | 0.73  [0.12-0.94] | 1.6 ± 50.8 | 5.2 ± 1.7  [3.0-7.7] | 0.51  [-0.13-0.88] | -29.1 ± 53.5 | 6.3 ± 3.0  [3.3-12.4] | 0.76  [0.22-0.95] | -14.2 ± 47.6 | 6.9 ± 2.1  [4.0-9.8] |
| ACA-L | 5.9 ± 3.0  [2.4-11.1] | 0.74  [0.28-0.93] | -4.3 ± 67.2 | 4.2 ± 1.4  [1.6-6.3] | 0.45  [-0.11-0.81] | -32.5 ± 69.2 | 5.7 ± 3.4  [2.1-13.4] | 0.78  [0.36-0.94] | -9.5 ± 66.8 | 5.8 ± 2.0  [2.8-10.2] |
| MCA-R | 10.7 ± 3.3  [3.2-15.9] | 0.50  [0.02-0.80] | 12.9 ± 58.0 | 8.7 ± 2.1  [4.1-11.8] | 0.73  [0.36-0.90] | -4.6 ± 41.8 | 9.7 ± 2.8  [4.0-16.3] | 0.62  [0.17-0.86] | 5.2 ± 47.2 | 9.1 ± 2.2  [4.8-13.2] |
| MCA-L | 9.6 ± 3.4  [3.5-15.3] | 0.63  [0.12-0.88] | 15.2 ± 49.4 | 7.0 ± 1.6  [3.9-9.1] | 0.35  [-0.17-0.74] | -11.5 ± 60.8 | 9.0 ± 2.6  [4.6-13.5] | 0.76  [0.36-0.92] | 11.1 ± 36.3 | 8.1 ± 2.5  [3.4-12.4] |
| PCA-R | 6.5 ± 2.4  [2.6-9.7] | 0.30  [-0.54-0.81] | 2.4 ± 77.8 | 4.9 ± 1.5  [2.6-7.7] | 0.31  [-0.23-0.79] | -22.4 ± 52.9 | 6.0 ± 1.9  [3.3-8.8] | 0.40  [-0.48-0.85] | 2.7 ± 65.3 | 6.1 ± 1.5  [3.7-7.9] |
| PCA-L | 7.2 ± 1.9  [4.4-9.7] | 0.29  [-0.30-0.78] | 14.0 ± 53.1 | 5.1 ± 1.5  [3.2-7.6] | 0.11  [-0.41-0.69] | -20.4 ± 59.0 | 6.8 ± 2.0  [3.4-9.8] | 0.19  [-0.53-0.76] | 8.8 ± 68.8 | 6.1 ± 1.1  [4.2-7.5] |
| ICA-R | 19.8 ± 4.7  [13.1-29.1] | 0.87  [0.68-0.95] | 5.7 ± 25.9 | 18.1 ± 5.8  [6.5-32.5] | 0.79  [0.53-0.92] | -5.9 ± 47.0 | 19.6 ± 4.5  [13.3-28.6] | 0.87  [0.67-0.95] | 5.2 ± 26.7 | 19.0 ± 5.9  [11.6-32.0] |
| ICA-L | 19.3 ± 5.7  [10.8-27.0] | 0.56  [0.11-0.82] | 8.5 ± 51.4 | 16.4 ± 2.6  [12.9-21.4] | 0.13  [-0.41-0.59] | -3.8 ± 49.0 | 18.9 ± 4.8  [12.2-28.4] | 0.68  [0.28-0.88] | 8.2 ± 34.9 | 17.3 ± 3.7  [11.3-22.4] |
| BA | 13.0 ± 3.3  [7.7-18.4] | 0.65  [-0.04-0.89] | 17.3 ± 29.2 | 9.7 ± 3.1  [3.6-13.7] | 0.61  [0.17-0.85] | -25.3 ± 52.7 | 11.3 ± 2.6  [7.0-16.5] | 0.76  [0.42-0.91] | 3.3 ± 35.9 | 10.9 ± 2.5  [6.3-14.3] |

***Supplementary Table 5:*** *Mean blood flow (mL/s) by artery: mean ± SD [range], intraclass correlation (ICC [95% CI]), and Bland-Altman bias ± limits of agreement (LOA, %) for three segmentation models vs. manual reference.*

|  | U-Net | | | DenseNet U-Net | | | Fine-tuned nnU-Net | | | Manual reference |
| --- | --- | --- | --- | --- | --- | --- | --- | --- | --- | --- |
|  | Mean ± SD [range] | ICC  [95% CI] | Bias ± LOA [%] | Mean ± SD [range] | ICC  [95% CI] | Bias ± LOA [%] | Mean ± SD [range] | ICC  [95% CI] | Bias ± LOA [%] | Mean ± SD [range] |
| ACA-R | 1.87 ± 1.09  [0.53-3.46] | 0.98  [0.90-1.00] | -2.69 ± 28.34 | 1.41 ± 0.72  [0.53-2.50] | 0.79  [0.07-0.96] | -23.65 ± 39.53 | 1.77 ± 1.10  [0.49-3.69] | 0.97  [0.85-0.99] | -9.32 ± 31.48 | 1.84 ± 0.94  [0.54-3.13] |
| ACA-L | 1.79 ± 0.99  [0.66-3.57] | 0.94  [0.81-0.99] | -5.92 ± 46.30 | 1.42 ± 0.61  [0.62-2.37] | 0.76  [0.08-0.94] | -23.92 ± 54.21 | 1.78 ± 1.04  [0.60-3.91] | 0.95  [0.81-0.99] | -7.64 ± 46.84 | 1.80 ± 0.80  [0.85-3.38] |
| MCA-R | 2.62 ± 1.20  [0.76-4.67] | 0.95  [0.85-0.99] | 2.53 ± 28.75 | 2.36 ± 0.81 [0.85-3.60] | 0.96  [0.88-0.99] | -3.20 ± 23.54 | 2.57 ± 1.15  [0.82-4.44] | 0.96  [0.88-0.99] | 2.00 ± 26.87 | 2.46 ± 0.96  [1.11-4.04] |
| MCA-L | 2.45 ± 1.20  [0.76-4.33] | 0.94  [0.78-0.98] | 6.62 ± 34.67 | 2.02 ± 0.64  [0.79-3.26] | 0.77  [0.40-0.93] | -5.26 ± 42.18 | 2.45 ± 1.13  [0.70-4.33] | 0.96  [0.76-0.99] | 8.44 ± 22.21 | 2.25 ± 1.02  [0.71-3.84] |
| PCA-R | 1.29 ± 0.55  [0.45-2.27] | 0.72  [0.09-0.94] | -0.07 ± 55.27 | 1.01 ± 0.28  [0.60-1.37] | 0.55  [-0.08-0.88] | -18.31 ± 39.53 | 1.23 ± 0.52  [0.45-2.06] | 0.78  [0.20-0.95] | -4.72± 51.21 | 1.22 ± 0.34  [0.73-1.71] |
| PCA-L | 1.50 ± 0.56  [0.96-2.48] | 0.82  [0.37-0.96] | 8.22 ± 33.08 | 1.15 ± 0.26  [0.80-1.67] | 0.54  [-0.08-0.88] | -14.77 ± 47.08 | 1.46 ± 0.56  [0.78-2.46] | 0.80  [0.34-0.96] | 4.45 ± 47.04 | 1.35 ± 0.37 [0.87-1.98] |
| ICA-R | 3.97 ± 1.53  [1.46-7.47] | 0.98  [0.90-0.99] | 4.76 ± 13.92 | 3.66 ± 1.36  [1.63-6.26] | 0.94  [0.83-0.98] | -2.86 ± 30.01 | 3.96 ± 1.47  [1.56-7.33] | 0.98  [0.91-0.99] | 4.91 ± 13.56 | 3.78 ± 1.42  [1.37-6.98] |
| ICA-L | 4.34 ± 1.43  [1.59-6.51] | 0.93  [0.82-0.98] | 1.63 ± 23.17 | 3.99 ± 1.03  [2.18-5.51] | 0.93  [0.78-0.98] | -4.07 ± 17.27 | 4.24 ± 1.38  [1.96-6.54] | 0.96  [0.89-0.99] | 0.04 ± 14.94 | 4.19 ± 1.20  [2.06-6.00] |
| BA | 2.37 ± 1.28  [0.78-4.58] | 0.98  [0.87-0.99] | 8.32 ± 23.27 | 1.93 ± 0.99  [0.53-3.40] | 0.88  [0.65-0.96] | -12.07 ± 40.04 | 2.22 ± 1.26  [0.70-4.57] | 0.98  [0.95-0.99] | 1.00 ± 27.38 | 2.20 ± 1.19  [0.71-4.26] |

***Supplementary Table 6:*** *Mean velocity (cm/s) by artery: mean ± SD [range], intraclass correlation (ICC [95% CI]), and Bland-Altman bias ± limits of agreement (LOA, %) for three segmentation models vs. manual reference.*

|  | U-Net | | | DenseNet U-Net | | | Fine-tuned nnU-Net | | | Manual reference |
| --- | --- | --- | --- | --- | --- | --- | --- | --- | --- | --- |
|  | Mean ± SD [range] | ICC  [95% CI] | Bias ± LOA [%] | Mean ± SD [range] | ICC  [95% CI] | Bias ± LOA [%] | Mean ± SD [range] | ICC  [95% CI] | Bias ± LOA [%] | Mean ± SD [range] |
| ACA-R | 23.9 ± 7.1 [12.5-31.1] | 0.87  [0.51-0.97] | -4.8 ± 27.8 | 27.7 ± 11.7 [10.5-43.5] | 0.92  [0.63-0.99] | 5.3 ± 30.2 | 26.2 ± 7.9 [13.0-32.9] | 0.91  [0.63-0.98] | 4.3 ± 26.5 | 25.3 ± 8.8 [13.6-37.4] |
| ACA-L | 30.1 ± 2.4 [26.4-34.9] | 0.48  [-0.15-0.83] | -1.1 ± 28.2 | 34.1 ± 7.5 [24.7-49.1] | 0.82  [0.11-0.96] | 9.5 ± 17.8 | 31.1 ± 2.8 [26.9-36.9] | 0.58  [-0.02-0.87] | 2.0 ± 26.1 | 30.9 ± 5.9 [24.4-43.9] |
| MCA-R | 24.1 ± 5.6 [12.5-33.0] | 0.69  [0.25-0.89] | -10.3 ± 36.3 | 28.2 ± 10.9 [13.5-47.5] | 0.97  [0.91-0.99] | 1.4 ± 20.0 | 26.1 ± 7.2 [15.6-39.9] | 0.92  [0.77-0.97] | -3.1 ± 22.7 | 27.5 ± 9.5 [14.9-43.7] |
| MCA-L | 25.4 ± 7.9 [11.1-36.3] | 0.82  [0.47-0.95] | -8.8 ± 29.4 | 30.2 ± 11.0 [10.6-47.2] | 0.92  [0.73-0.98] | 6.7 ± 27.8 | 26.9 ± 8.0 [10.1-39.5] | 0.93  [0.79-0.98] | -2.8 ± 25.3 | 28.1 ± 9.6 [11.0-41.8] |
| PCA-R | 20.1 ± 5.4 [10.9-27.6] | 0.86  [0.48-0.97] | -2.2 ± 26.9 | 21.7 ± 7.1 [13.9-32.0] | 0.94  [0.75-0.99] | 4.3 ± 27.8 | 20.4 ± 5.8 [12.4-28.0] | 0.90  [0.57-0.98] | -1.0 ± 27.6 | 20.8 ± 6.9 [11.2-31.8] |
| PCA-L | 21.0 ± 4.1 [14.9-25.5] | 0.84  [0.42-0.97] | -5.7 ± 21.8 | 23.9 ± 6.0 [15.6-31.0] | 0.91  [0.60-0.98] | 6.1 ± 20.2 | 21.3 ± 3.4 [15.9-25.3] | 0.76  [0.24-0.95] | -3.7 ± 29.0 | 22.5 ± 5.7 [14.7-29.4] |
| ICA-R | 20.0 ± 5.6 [9.3-28.9] | 0.97  [0.91-0.99] | -0.9 ± 15.1 | 21.4 ±7.9 [9.0-33.3] | 0.92  [0.81-0.97] | 3.2 ± 23.7 | 20.3 ± 6.5 [8.9-31.5] | 0.97  [0.91-0.99] | -0.2 ± 16.9 | 20.5 ± 6.9 [8.9-32.5] |
| ICA-L | 23.1 ± 6.9 [14.0-39.9] | 0.79  [0.48-0.92] | -7.0 ± 34.3 | 24.8 ± 7.2 [12.1-36.0] | 0.80  [0.49-0.93] | -0.1 ± 36.1 | 23.2 ± 6.8 [11.3-35.6] | 0.83  [0.56-0.94] | -7.0 ± 29.2 | 25.1 ± 8.1 [12.5-38.6] |
| BA | 17.4 ± 6.9 [6.8-34.5] | 0.94  [0.55-0.98] | -9.4 ± 23.1 | 20.6 ± 10.7 [6.3-50.5] | 0.93  [0.81-0.98] | 3.6 ± 27.8 | 18.8 ± 7.8 [7.3-37.0] | 0.98  [0.93-0.99] | -2.6 ± 21.2 | 19.4 ± 8.1 [8.1-38.3] |

***Supplementary Table 7****: Mean and max wall shear stress (WSS, Pa): mean ± SD [range], intraclass correlation (ICC [95% CI]), and Bland-Altman bias ± limits of agreement (LOA, %) for three segmentation models vs. manual reference.*

|  | U-Net | | | DenseNet U-Net | | | Fine-tuned nnU-Net | | | GT |
| --- | --- | --- | --- | --- | --- | --- | --- | --- | --- | --- |
|  | Mean ± SD [range] | ICC  [95% CI] | Bias ± LOA [%] | Mean ± SD [range] | ICC  [95% CI] | Bias ± LOA [%] | Mean ± SD [range] | ICC  [95% CI] | Bias ± LOA [%] | Mean ± SD [range] |
| Mean WSS | 1.44 ± 0.53 [0.56-2.25] | 0.95  [0.85-0.98] | -5.21 ± 16.55 | 1.64 ± 0.66 [0.75-3.14] | 0.93  [0.79-0.97] | 7.27 ± 22.91 | 1.57 ± 0.63 [0.55-2.52] | 0.96  [0.89-0.98] | 1.69 ± 17.65 | 1.53 ± 0.59 [0.61-2.82] |
| Max WSS | 1.99 ± 0.91 [0.64-3.31] | 0.97  [0.92-0.99] | -4.76 ± 15.39 | 2.23 ± 1.07 [0.90-4.11] | 0.96  [0.87-0.98] | 6.89 ± 20.93 | 2.16 ± 1.05 [0.62-3.63] | 0.97  [0.93-0.99] | 1.35 ± 16.94 | 2.09 ± 0.98 [0.71-3.88] |

***Supplementary Table 8****: Segmentation performance of DenseNet U-Net trained for 100 and 300 epochs. All values are presented as mean [range].*

|  | | **100 epochs** | | **300 epochs** | |
| --- | --- | --- | --- | --- | --- |
|  |  | **Run 1** | **Run 2** | **Run 1** | **Run 2** |
| Dice score | Low resolution | 0.813  [0.649-0.919] | 0.801  [0.609-0.917] | 0.823  [0.691-0.923] | 0.799  [0.591-0.923] |
|  | High resolution | 0.732  [0.270-0.869] | 0.706  [0.226-0.863] | 0.757  [0.475-0.879] | 0.687  [0.189-0.865] |
| HD95 (mm) | Low resolution | 5.06  [0.66-14.07] | 5.36  [0.66-15.80] | 4.59  [0.66-24.96] | 5.55  [0.66-21.83] |
|  | High resolution | 4.95  [0.50-22.94] | 6.66  [0.50-28.66] | 5.55  [0.66-21.83] | 7.41  [0.50-28.94] |

***Supplementary Figure 1:*** *Analysis plane placement for hemodynamic measurements in the Circle of Willis (CoW).* *ACA, anterior cerebral artery; BA, basilar artery; ICA, internal carotid artery; MCA, middle cerebral artery; PCA, posterior cerebral artery. L and R denote left and right, respectively.*
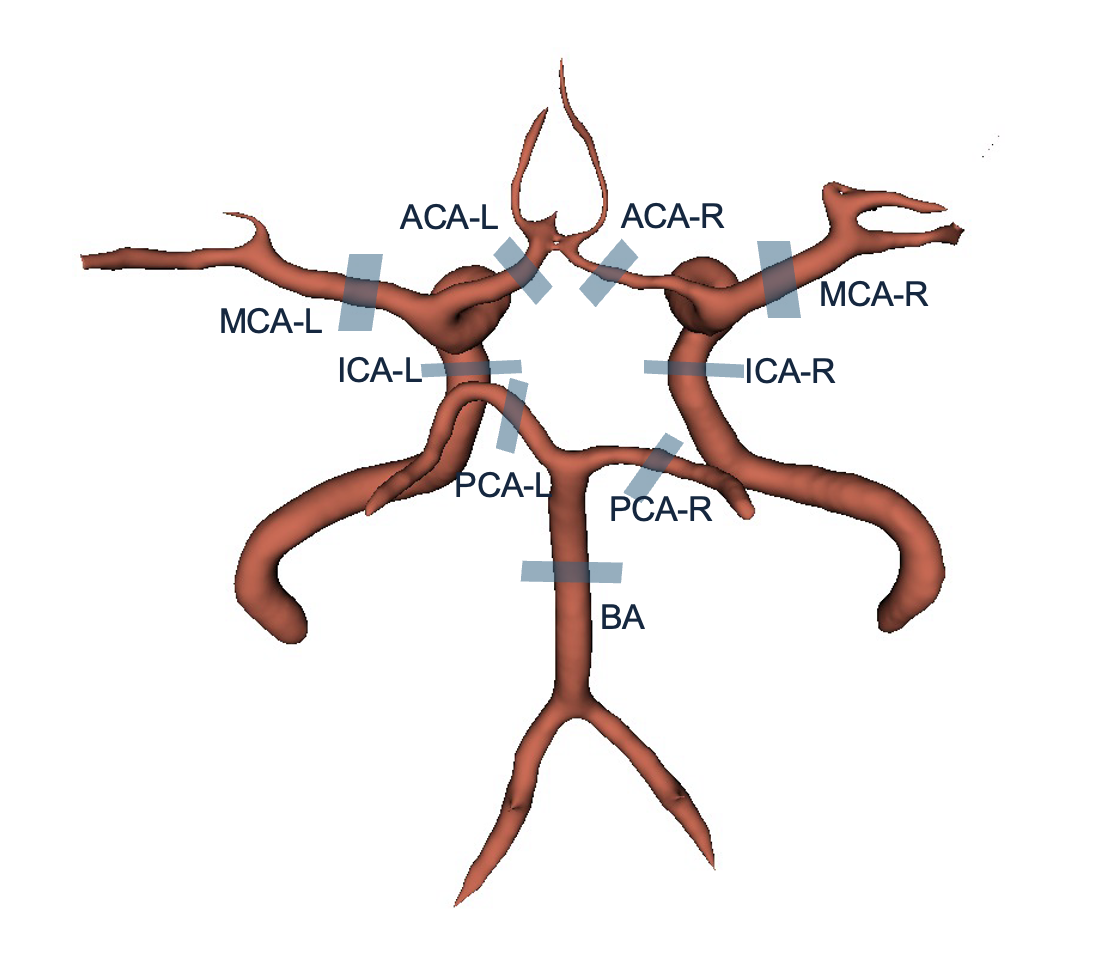

***Supplementary Figure 2:*** *Effect of 4D Flow MRI training-set size on fine-tuned and from-scratch nnU-Net performance. Dice score and HD95 are shown for the low- and high-resolution test sets. The fine-tuned nnU-Net was compared with from-scratch nnU-Net models trained for 200 and 1000 epochs using reduced training sets of 1, 3, 5, 7, 9, and 11 cases.*

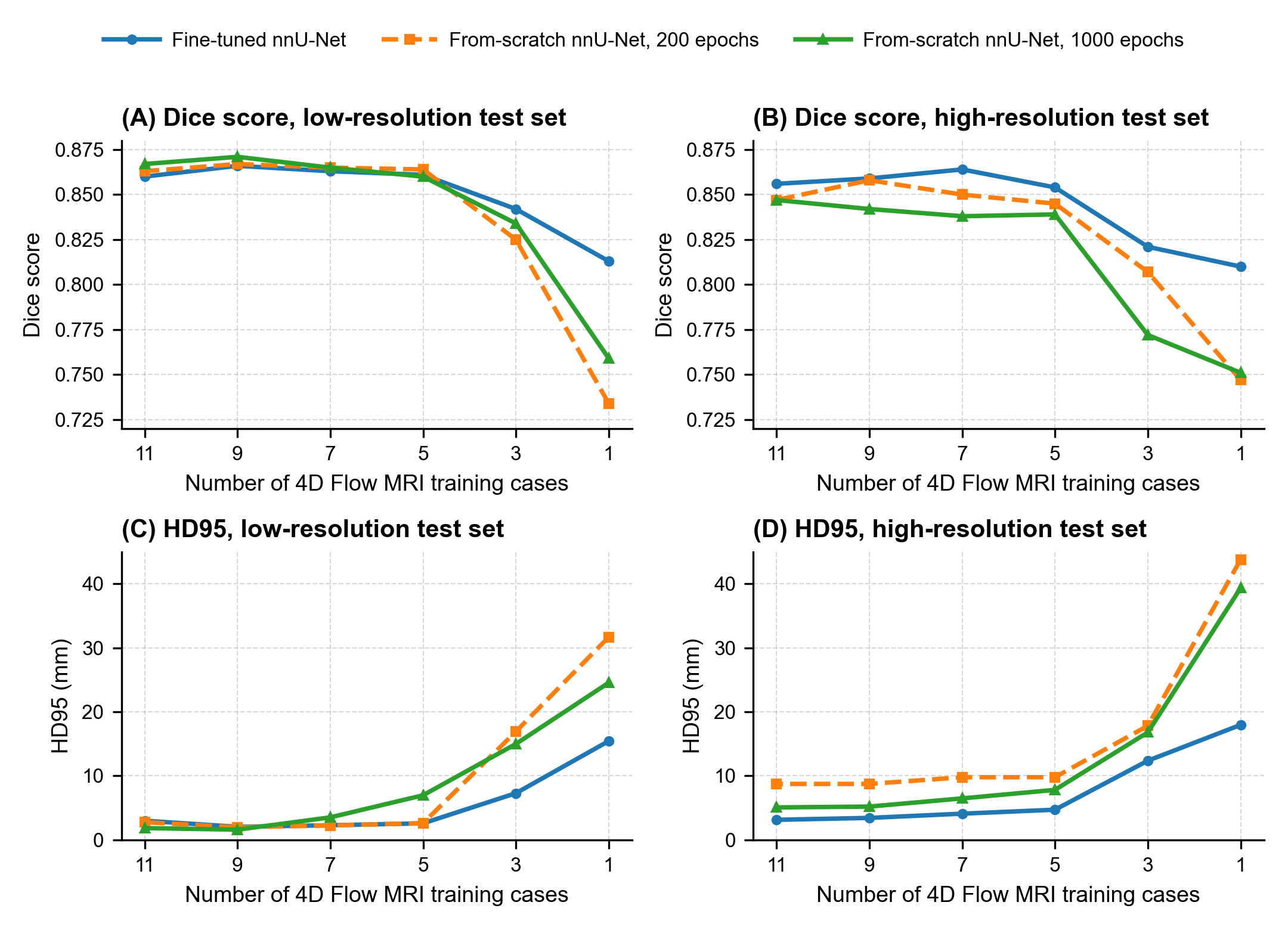
